# Chronic kidney disease is associated with attenuated plasma metabolome response to oral glucose tolerance testing

**DOI:** 10.1101/2022.01.08.22268946

**Authors:** Armin Ahmadi, M. Nazmul Huda, Brian J. Bennett, Jorge Gamboa, Leila R. Zelnick, Lucas R. Smith, Maria Chondronikola, Daniel Raftery, Ian H. de Boer, Baback Roshanravan

## Abstract

Chronic kidney disease (CKD), a major public health problem, is associated with decreased anabolic response to insulin contributing to protein-energy wasting. Targeted metabolic profiling of the response to oral glucose tolerance testing (OGTT) may help identify metabolic pathways contributing to disruptions to insulin response in CKD. Using targeted metabolic profiling, we examined plasma metabolome in 41 moderate-to-severe non-diabetic CKD patients with estimated glomerular filtration rate (eGFR)<60ml/min per 1.73m^2^ (38.9±12.7) and 20 healthy controls with normal eGFR (87.2±17.7) before and after 2h of 75g oral glucose load. Compared to controls, CKD participants had higher lactate: pyruvate (L:P) ratio both at fasting and after oral glucose challenge. Total energy production estimated through GTP:GDP ratio was impaired during OGTT despite similar fasting GTP:GDP ratio. CKD group had sustained elevation of vitamin B family members, TCA cycle metabolites, and purine nucleotides in response to glucose challenge. Metabolic profiling in response to OGTT suggests a broad disruption of mitochondrial energy metabolism in CKD patients. These findings motivate further investigation into insulin sensitizers in patients with non-diabetic CKD and their impact on energy metabolism.

## Introduction

Chronic kidney disease (CKD) has been recognized as a public health burden worldwide with the estimated global prevalence of 14% affecting over 650 million people globally [1]. Insulin resistance (IR) is one of the very early metabolic alterations in CKD increasing with severity of kidney disease [2]. IR is considered a cardio-metabolic risk factor correlated with increased systemic inflammation [3] and oxidative stress [4]. Furthermore, impaired glucose tolerance has been closely associated with increased risk of all-cause and cardiovascular mortality [5, 6].

Multiple pathophysiologic features of CKD contribute to IR in CKD [7] including chronic inflammation [8–10], altered gut microbiome [11], oxidative stress [12–14], metabolic acidosis [15], and accumulation of uremic toxins [11]. However, evidence for the biologic basis for IR in humans with CKD is lacking.

Studies in animal models indicate the IR related to CKD is the result of changes in insulin signaling both at the receptor and post-receptor levels [16] leading to blunted glucose uptake, metabolism, and storage in target tissues [17]. Peripheral tissues such as skeletal muscle are the main sites impacted by IR in CKD and link these early metabolic alterations with functional decline and frailty [18]. Compromised mitochondrial bioenergetics and physical performance has been reported in patients with CKD [19]. Post-receptor defects observed in CKD may be the consequence of excess insulin stimulation and heightened inflammation leading to augmented degradation of the insulin receptor by the ubiquitin proteosome pathway [20].

Although tissue insensitivity to insulin is the main contributor to IR, changes in insulin secretion and degradation have also been established in CKD [21]. In particular, a recent study using the gold-standard high-dose hyperinsulinemic-euglycemic clamp to investigate peripheral insulin sensitivity showed disturbances in insulin clearance as a principle characteristic distinguishing patients with non-diabetic CKD to controls [22]. This study demonstrated profound alterations in the plasma metabolome response to insulin in patients with CKD compared to controls.

However, understanding of the biologic basis for IR in humans with CKD at more physiologic, endogenous levels of insulin, such as that elicited by oral glucose tolerance testing (OGTT) remain unknown.

Oral glucose tolerance tests provide estimates of IR [23] but also captures additional physiologic processes by stimulating gut-derived incretin hormones known to augment insulin secretion [24, 25]. Metabolomic profiling of the response to OGTT may reveal specific metabolites and metabolic pathways underlying impaired glucose homeostasis in CKD thus identifying therapeutic targets for improving insulin sensitivity in this vulnerable population. We performed targeted plasma metabolic profiling comparing non-diabetic patients with moderate-severe CKD to healthy controls during an oral glucose challenge. In this study, we evaluated if CKD is associated with impaired plasma metabolic response to oral glucose challenge compared to controls. Our goal was to identify specific metabolic alterations in non-diabetic CKD to better understand the metabolic and biological basis for glucose intolerance and IR in CKD.

## Results

### Characteristics of the study participants

The study included 41 CKD participants with a mean eGFR of 38.9±12.7 ml/min per 1.73 m^2^, and 20 control subjects with a mean eGFR of 87.2±17.7 ml/min per 1.73 m^2^. The mean age was 61.2±12.9 years, 44% were women, and 22% were self-reported Black (**Table 1**). Compared with controls, participants with CKD had higher body weight, lower fat-free mass, higher BMI, and daily calorie intake, and greater plasma inflammatory markers.

**Table 1.** Demographic characteristics by CKD status of analytic population (n=61). Abbreviations: Chronic kidney disease was defined as estimated glomerular filtration rate <60 ml/min per 1.73 m^2^; controls as ≥60 ml/min per 1.73 m^2^. SD, standard deviation; IQR, interquartile range; CCB, calcium channel blocker; RAASi, renin-angiotensin-aldosterone system inhibitor; eGFR, estimated glomerular filtration rate; CRP, C-reactive protein.

|  | **Controls**<br>N=20 | **Chronic Kidney disease**<br>N=41 |
| --- | --- | --- |
| **Demographics:** |  |  |
| Age (yr), mean (SD) | 60.3 (12.9) | 61.6 (14.3) |
| Female, number (%) | 9 (45) | 18 (44) |
| **Race/ethnicity, number (%)** |  |  |
| White | 16 (80) | 28 (68) |
| Black | 4 (20) | 10 (24) |
| Asian/Pacific Islander | 0 (0) | 3 (7) |
| **Physical characteristics, mean (SD)** |  |  |
| Weight (Kg) | 85.9(21.4) | 89.5(20.3) |
| Fat-free mass (kg) | 54.3(11.9) | 51.9(11.5) |
| BMI, mean (SD) | 28.7(6.43) | 30.2(6.16) |
| **Dietary data** |  |  |
| Total calorie intake (kcal), mean (SD) | 1987.1(615.3) | 1819.0(573.1) |
| Daily protein intake (g), mean (SD) | 75.0(23.1) | 74.4(29.8) |
| **Laboratory data:** |  |  |
| eGFR (mL/min/1.73m<sup>2</sup>), mean (SD) | 87.2 (17.7) | 38.9 (12.7) |
| Serum creatinine (mg/dL), median (IQR) | 0.9 (0.81-1.0) | 1.67 (1.50-1.80) |
| Serum cystatin C (mh/dL), median (IQR) | 0.89 (0.68-0.97) | 1.49 (1.40-1.81) |
| Urine albumin excretion rate (mg/24 hours), median (IQR) | 5 (3.4-7.8) | 127.15 (20.57-422.85) |
| CRP (mg/dL), median (IQR) | 0.11 (0.05-0.25) | 0.23 (0.13-0.27) |
| IL-1β (pg/mL), median (IQR) | 0.09 (0.05-0.14) | 0.12 (0.08-0.15) |
| IL-6 (pg/mL), median (IQR) | 0.84 (0.56-1.19) | 0.99 (0.83-1.51) |
| IFN-γ (pg/mL), median (IQR) | 2.24 (1.09-4.15) | 3.26 (2.59-5.62) |
| TNFα (pg/mL), median (IQR) | 1.49 (0.75-1.95) | 2.53 (2.23-2.85) |
| **Baseline medication:** |  |  |
| Antihypertensive medication, number (%) | 8 (40.0) | 31 (86.1) |
| RAASi | 4 (20) | 22 (61.11) |
| Diuretic | 2 (10) | 15 (41.6) |
| Statin | 3 (15) | 14 (38.9) |
| Beta blocker | 2 (10) | 12 (33.3) |
| CCBs | 2(10) | 18 (50) |

Oral glucose challenge (OGTT) was associated with a significant reduction in a wide range of metabolites primarily amino acids, purine nucleotides, and dicarboxylic acids. After adjusting for age, sex, race/ethnicity, body weight, and LC-MS batch, 65% (58/88) of the detected plasma metabolites were significantly altered post oral glucose challenge in the overall cohort (**Table 2** and **Supplemental Table 1**). The largest reductions in individual metabolite concentrations post OGTT were observed for linoleic acid (fatty acid metabolism), ADP (nucleotide/purine nucleotide), and nicotinamide (vitamin) with a percent change of -82%, -79%, and -63%, respectively (**Table 2**). Only 6% (5/88) of the detected metabolites significantly increased post OGTT compared to fasting state. These metabolites included erythrose (sugar), glucose (glycolysis/sugar), and kynurenate (tryptophan cycle) with a percent change of +53%, +28%, and +17% respectively (**Supplemental Table 2**).

**Table 2.** Differences in plasma metabolites during OGTT, compared with the fasting state

| **Metabolite** | **Fold change between the OGTT/fasting states (95% CI)** | **p-value** |
| --- | --- | --- |
| Linolenic acid | 0.18 (0.15, 0.21) | < 0.001 |
| ADP | 0.21 (0.14, 0.32) | < 0.001 |
| Niacinamide | 0.37 (0.29, 0.48) | < 0.001 |
| Malonic acid | 0.38 (0.28, 0.51) | < 0.001 |
| Taurine | 0.49 (0.41, 0.58) | < 0.001 |
| Aspartic acid | 0.51 (0.43, 0.6) | < 0.001 |
| Adenylosuccinate | 0.53 (0.44, 0.65) | < 0.001 |
| iso-Leucine | 0.55 (0.5, 0.6) | < 0.001 |
| Leucine | 0.56 (0.51, 0.61) | < 0.001 |
| Citrulline | 0.58 (0.53, 0.62) | < 0.001 |
| PGE | 0.6 (0.53, 0.69) | < 0.001 |
| Succinate | 0.61 (0.55, 0.68) | < 0.001 |
| Tyrosine | 0.65 (0.61, 0.69) | < 0.001 |
| Sorbitol | 0.66 (0.6, 0.72) | < 0.001 |
| Methionine | 0.67 (0.62, 0.71) | < 0.001 |
| Cystathionine | 0.7 (0.59, 0.84) | < 0.001 |
| Glutamic acid | 0.7 (0.6, 0.82) | < 0.001 |
| 2-Hydroxyglutarate | 0.7 (0.62, 0.78) | < 0.001 |
| Serine | 0.71 (0.66, 0.76) | < 0.001 |
| MethylSuccinate | 0.72 (0.67, 0.78) | < 0.001 |

Top 20 based on p-value and fold change are shown in the table. Fold changes are adjusted to age, sex, race, body weight and batch effect. The fold changes are compared to the fasting state. For example, Nicotinamide has fold change of 0.37 meaning it had a 63% reduction post OGTT compared to the fasting state.

CKD attenuates the plasma metabolome response to glucose challenge in metabolites from predominantly the vitamin B family, TCA cycle intermediates, and purine nucleotides. After adjustment, changes in 15% (13/88) of detected plasma metabolites were significantly altered between CKD and controls post oral glucose challenge (**Table 3** and **Supplemental Table 2**). Overall, CKD was associated with higher plasma levels of these metabolites such as succinate, inositol, nicotinamide, and glucose in response to OGTT compared to control. A notable exception to this was kynurenate. In general, controls had on average a greater decline in plasma metabolites in response to the oral glucose challenge compared to participants with CKD. The largest difference in response to the glucose challenge comparing CKD and controls were observed in ADP (nucleotide/purine nucleotide), glycochenodeoxycholate (bile acid metabolism), and IMP (nucleotide/purine metabolism). The ratio in fold change comparing CKD to control for these metabolites were 3.7, 3.6, and 3.1 respectively (**Table 3**).

**Table 3.**
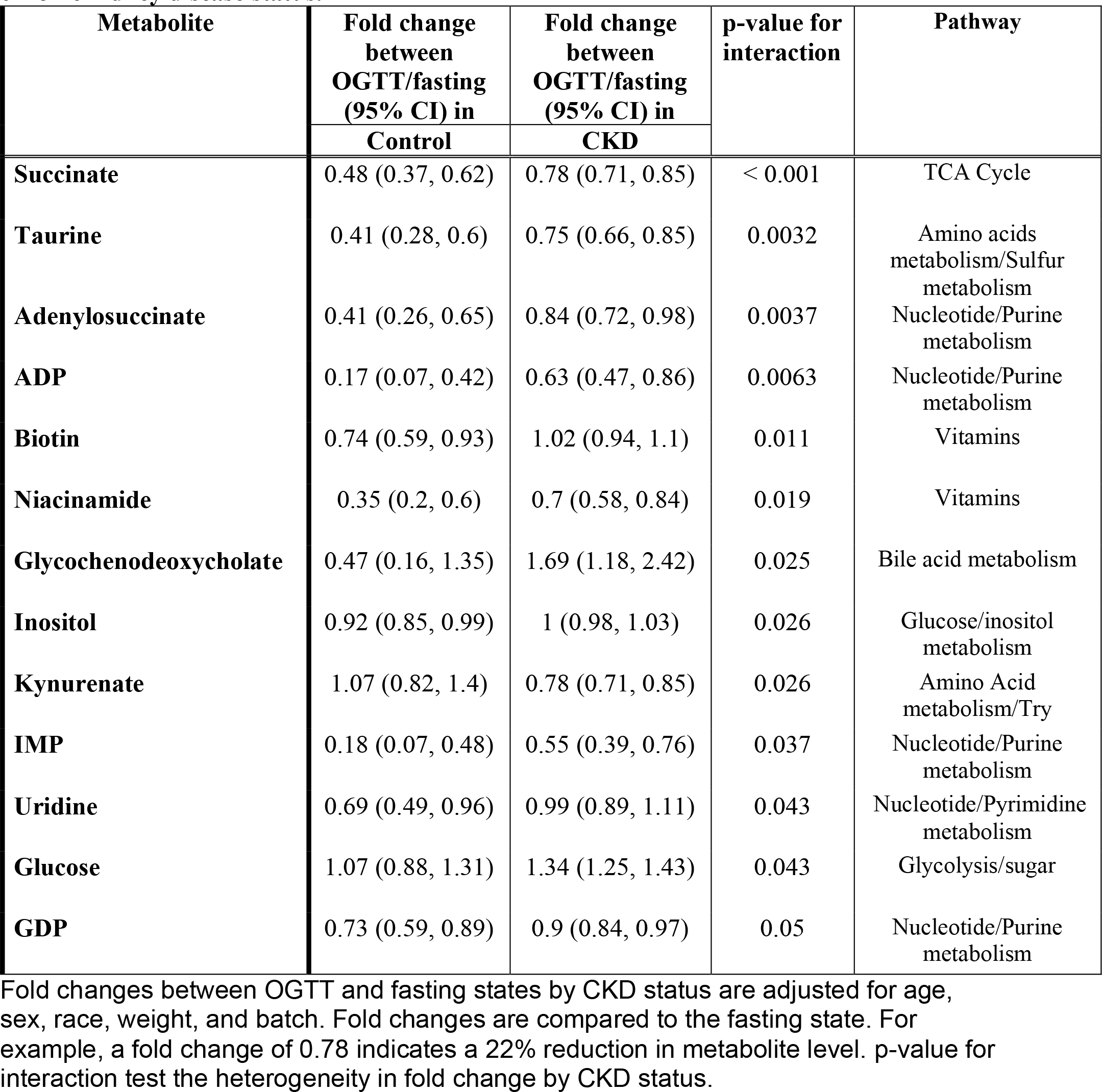
Differences in plasma metabolites response post oral glucose tolerance test by chronic kidney disease status.

| Metabolite | Fold change between OGTT/fasting (95% CI) in | Fold change between OGTT/fasting (95% CI) in | p-value for interaction | Pathway |
| --- | --- | --- | --- | --- |
|  | Control | CKD |  |  |
| **Succinate** | 0.48 (0.37, 0.62) | 0.78 (0.71, 0.85) | < 0.001 | TCA Cycle |
| **Taurine** | 0.41 (0.28, 0.6) | 0.75 (0.66, 0.85) | 0.0032 | Amino acids metabolism/Sulfur metabolism |
| **Adenylosuccinate** | 0.41 (0.26, 0.65) | 0.84 (0.72, 0.98) | 0.0037 | Nucleotide/Purine metabolism |
| **ADP** | 0.17 (0.07, 0.42) | 0.63 (0.47, 0.86) | 0.0063 | Nucleotide/Purine metabolism |
| **Biotin** | 0.74 (0.59, 0.93) | 1.02 (0.94, 1.1) | 0.011 | Vitamins |
| **Niacinamide** | 0.35 (0.2, 0.6) | 0.7 (0.58, 0.84) | 0.019 | Vitamins |
| **Glycochenodeoxycholate** | 0.47 (0.16, 1.35) | 1.69 (1.18, 2.42) | 0.025 | Bile acid metabolism |
| **Inositol** | 0.92 (0.85, 0.99) | 1 (0.98, 1.03) | 0.026 | Glucose/inositol metabolism |
| **Kynurenate** | 1.07 (0.82, 1.4) | 0.78 (0.71, 0.85) | 0.026 | Amino Acid metabolism/Try |
| **IMP** | 0.18 (0.07, 0.48) | 0.55 (0.39, 0.76) | 0.037 | Nucleotide/Purine metabolism |
| **Uridine** | 0.69 (0.49, 0.96) | 0.99 (0.89, 1.11) | 0.043 | Nucleotide/Pyrimidine metabolism |
| **Glucose** | 1.07 (0.88, 1.31) | 1.34 (1.25, 1.43) | 0.043 | Glycolysis/sugar |
| **GDP** | 0.73 (0.59, 0.89) | 0.9 (0.84, 0.97) | 0.05 | Nucleotide/Purine metabolism |

Fold changes between OGTT and fasting states by CKD status are adjusted for age, sex, race, weight, and batch. Fold changes are compared to the fasting state. For example, a fold change of 0.78 indicates a 22% reduction in metabolite level. p-value for interaction test the heterogeneity in fold change by CKD status.

The response to the oral glucose challenge demonstrates a broad disruption of amino acid and mitochondrial energy metabolism in CKD patients. In the entire cohort, oral glucose challenge impacted alpha-Linoleic acid metabolism (p-value=3.84 x 10^-31^), aminoacyl-tRNA biosynthesis (p-value=1.15 x 10^-26^) and arginine biosynthesis (p-value=2.66 x 10^-25^) (**Figure 1A**). In addition, metabolic pathways involving amino acid metabolism such as branch chain amino acids (valine, leucine, isoleucine), phenylalanine, and histidine metabolism were also impacted. Compared to controls, participants with CKD demonstrated significant aberrations in alpha-Linoleic acid metabolism (p-value=2.91 x 10^-5^), nicotinamide metabolism (p-value=5.18 x 10^-5^), and arginine biosynthesis (p-value=1.96 x 10^-4^) (**Figure 1B**).

**Figure 1.**
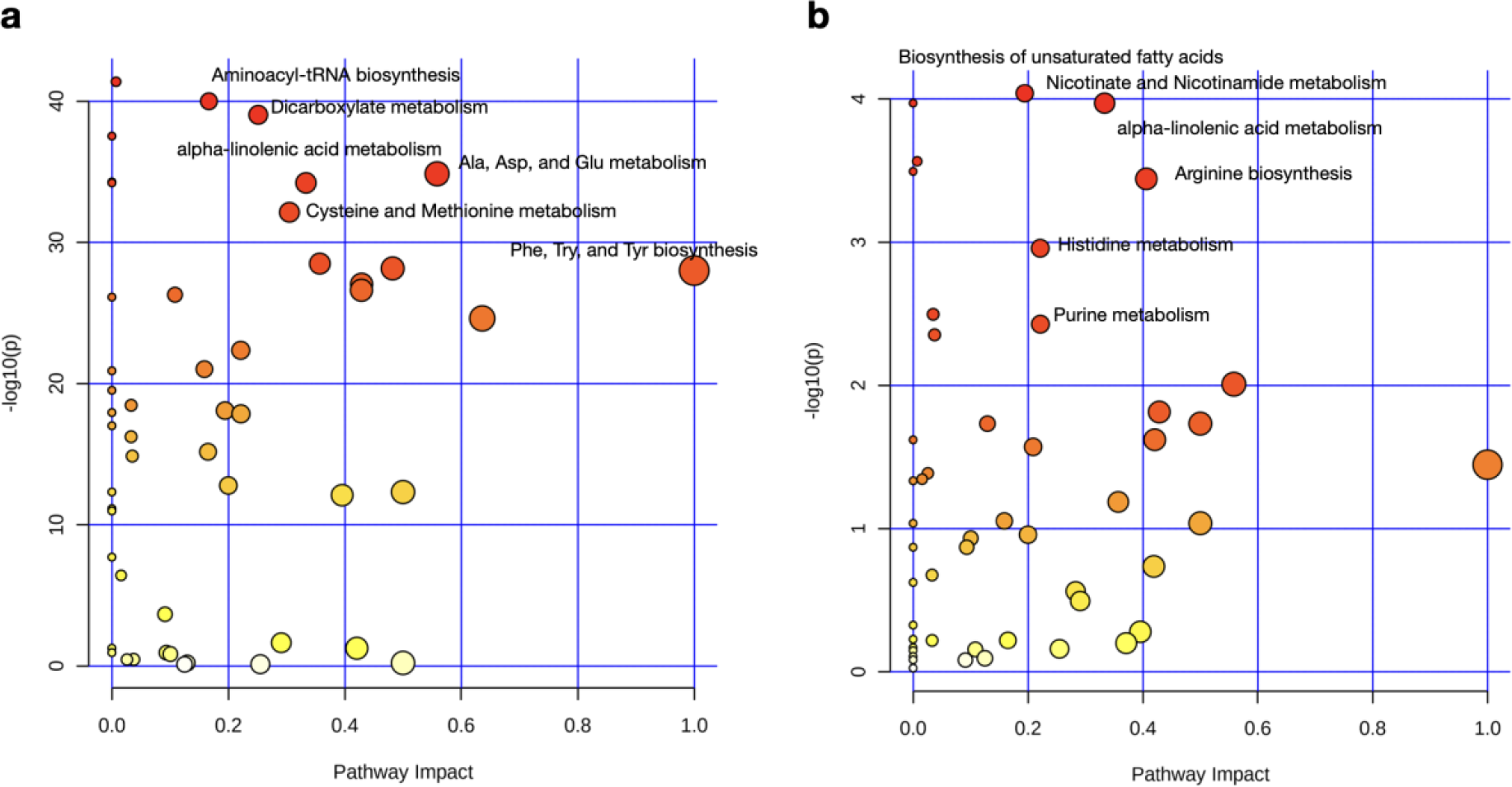
Pathway analysis of changes with oral glucose tolerance test. (**A**) in the overall cohort (n=61) and (**B**) comparing CKD (n=41) with controls (n=20). The size and color of the nodes represent pathway impact value and p-value, respectively.

### CKD is associated with greater lactate to pyruvate (L:P) ratio, an indicator of mitochondrial respiratory chain impairment

A lactate to pyruvate ratio greater than 30 in patients suggests a primary respiratory chain dysfunction [16]. L:P ratio at Fasting and post-OGTT was significantly higher in participants with CKD compared to controls (**Figure 2A**). The mean L:P ratio at fasting was 28.9 (IQR of 26.2, 31.2) for controls compared to 36.7 (IQR of 29.3, 42.3) among participants with CKD (p-value=0.0012). The high L:P ratio was sustained post OGTT with a mean of 32.8 (IQR of 30.7, 37.2) in CKD in contrast to 29.5 (IQR of 27.8, 32.7) in controls (p-value<0.0023) (**Figure 2A**). The difference in L:P ratio is mostly driven by an increase in pyruvate levels however, lactate (fold change (FC), of 1.15 vs 1.20) and pyruvate (FC of 1.08 vs 1.34) elevations were not meaningfully different in CKD compared to controls during OGTT **(Supplemental Table 2**).

**Figure 2.**
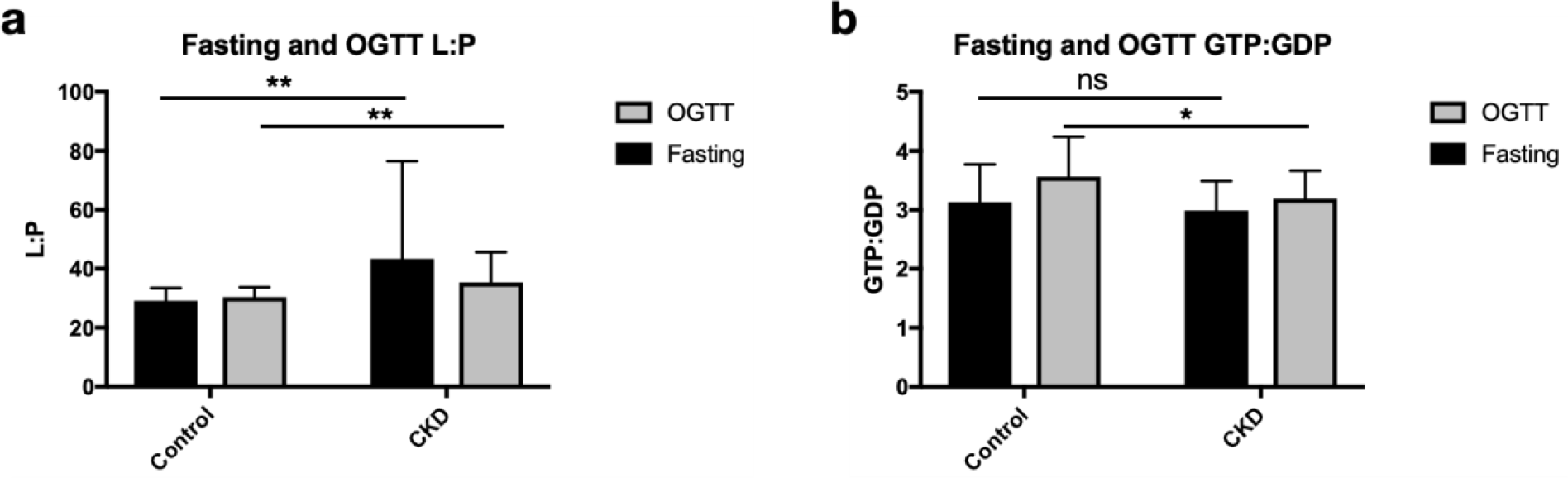
Distribution of L:P and GTP:GDP ratio in persons with CKD (n=41) and controls (n=21). Panel A: L:P ratio at fasting and OGTT comparing CKD to controls. Panel B: GTP:GDP ratio at fasting and during OGTT comparing CKD to controls. Data points represent mean and error bars represent SD. Statistical analysis using Mann Whitney 2 tailed test *p-value<0.05, **p-value<0.01.

Persons with CKD have similar global energy production as the controls indicated by equivalent GTP:GDP ratio but an impaired response to OGTT. To assess the level of global energy production and the changes after the glucose challenge, we looked at the GTP:GDP ratio during fasting and OGTT separately. The GTP:GDP ratio is known to reflect the ATP:ADP ratio [26]. After the oral glucose challenge, it is expected that the GTP and ATP levels will increase and ADP and GDP levels will decrease, thus resulting in a greater GTP:GDP and ATP:ADP ratios.

The median GTP:GDP ratio at fasting was similar in CKD and controls (p-value=0.056) (**Figure 2B**). As expected, the GTP:GDP ratio increased during OGTT compared to fasting in both groups with the median of 3.18 to 3.47 in controls (p-value=0.042) and 2.97 to 3.24 in CKD (p-value=0.007); however, the ratio did not increase as sharply with the median of 3.47(IQR of 3.05, 3.95) and 3.24 (IQR of 2.824, 3.48) in controls and CKD respectively (p-value=0.044) (**Figure 2B**). This difference is more evident when considering GTP:GDP ratio of each group separately. Similar fasting GTP:GDP ratios and lower GTP:GDP ratio at OGTT suggest an impaired insulin response in persons with CKD.

### Insulin secretion and sensitivity measured during OGTT did not differ among participants with and without CKD

We evaluated plasma insulin concentrations measured at 0, 10, 20, 30, 60, 90, and 120 minutes after glucose ingestion and found no significant differences between CKD and controls (**Figure 3A**). Similarly, the Matsuda index estimation of insulin sensitivity was not significantly different between CKD compared to controls (p-value=0.08) (**Figure 3B**). The median Matsuda index in CKD and controls was 3.93 (IQR of 2.44, 5.63) and 3.70 (IQR of 2.30, 9.13) respectively.

**Figure 3.**
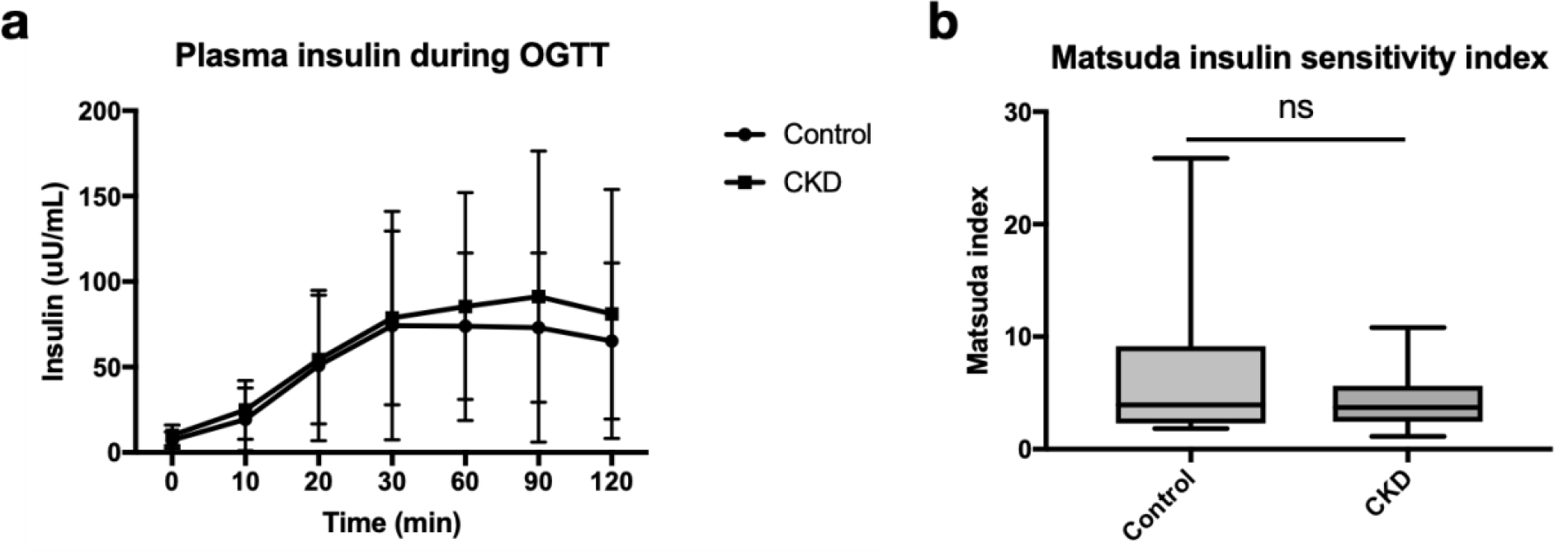
Measurements of insulin secretion and insulin sensitivity comparing CKD(n=40) and controls(n=20) during OGTT. A) Plasma insulin concentration. Data points represent means and error bars represent 95% CI. B) Matsuda index.

Changes in plasma metabolites in response to the oral glucose challenge is associated with CKD signature and inflammation markers. We performed weighted gene co-expression network analysis (WGCNA) to better understand how closely related metabolite groups associate with CKD. We identified 8 metabolite modules in response to glucose load. Of the total 8 metabolite modules, those indicated in blue and green modules were significantly correlated with CKD signature and known CKD-associated inflammation markers (**Figure 4A, 4B, and 4C**). Green module positively correlated with CKD status, plasma cystatin C, and plasma creatinine and negatively correlated with eGFR (**Figure 4B**). Some notable metabolites from the green module include uric acid, kynurenate, and malonic acid (**Supplemental Table 3**). The blue module also positively correlated with plasma cystatin C and plasma creatinine (**Figure 4C**). Both modules also positively correlated with plasma TNF-α (**Figure 4C**). Blue module predominantly consisted of amino acids such as serine, phenylalanine, asparagine, and arginine (**Supplemental Table 3**).

**Figure 4:**
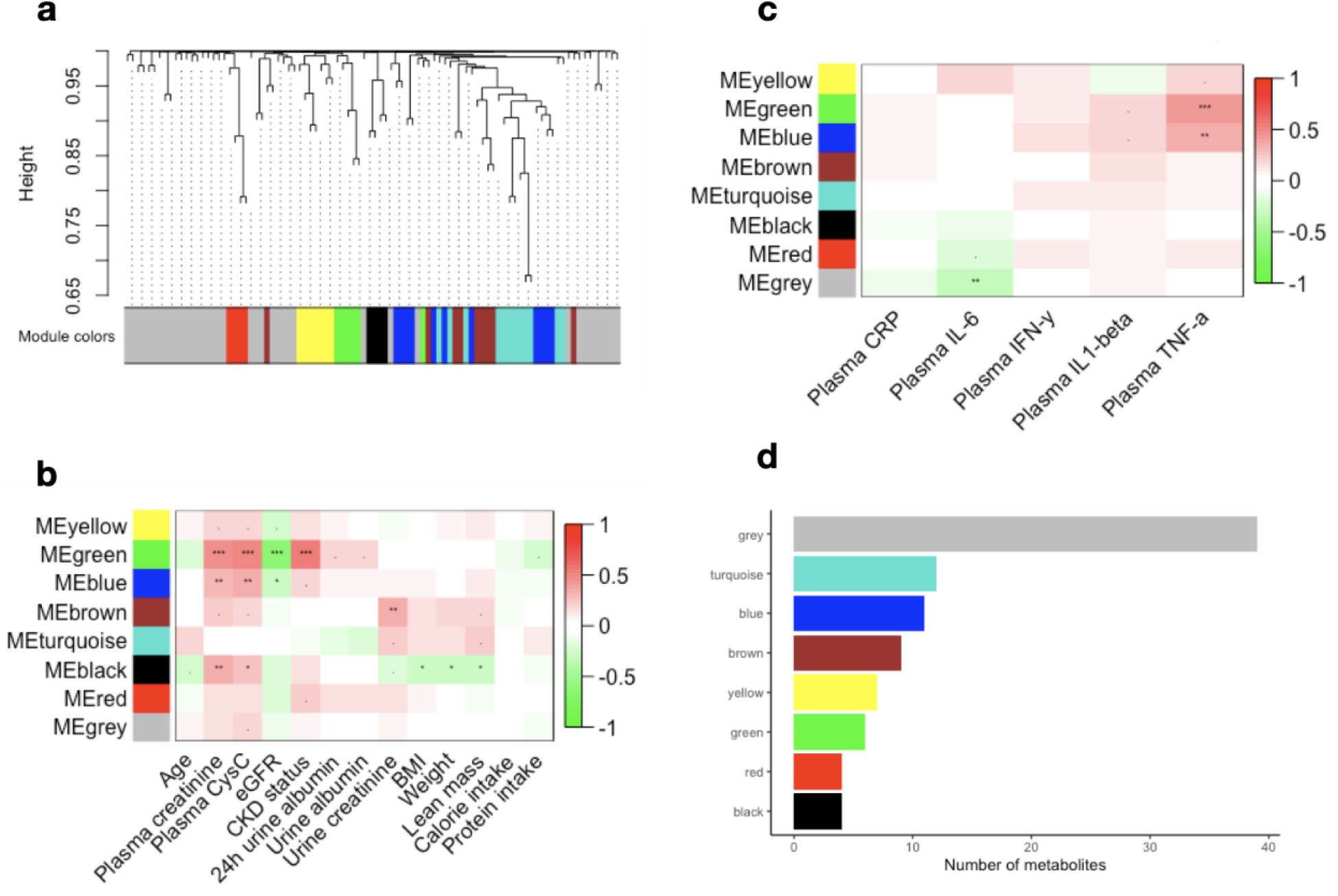
Changes in the plasma metabolic profile due to an oral glucose load is associated with kidney disease and inflammatory markers. a) Identified metabolite modules. Correlation between plasma metabolite modules and b) plasma biomarkers of kidney disease and c) inflammation markers. Each row in the table corresponds to a module, and each column represents a kidney biomarker, as indicated. The correlation coefficient is color-coded as indicated in the color key legend (red = positive and green = negative correlation) “***” = p-value<0.001, “**” = p-value<0.01, “*” = p-value<0.05, “.” = p-value<0.10. (d) The number of metabolites in each of the metabolite-module.

## Discussion

Using targeted metabolic profiling, we identified several attenuated biological and metabolic pathways in response to an oral glucose challenge in the overall cohort with evidence of marked heterogeneity by CKD status. First, we observed that compared to the fasting state OGTT resulted in a significant decrease in a wide range of metabolites including purine nucleotides and amino acids in the overall cohort. Second, patients with CKD demonstrate an attenuated plasma metabolome response to oral glucose challenge. Our prior findings in this cohort were confirmed that these changes occur in the absence of significant differences in insulin secretion and insulin sensitivity (**Figure 3A and 3B**) [22]. In response to the glucose challenge, we observed changes in 13 metabolites significantly differ in participants with CKD compared to controls, a significant portion of which are known uremic toxins associated in persons with CKD. Third, we observed an attenuated plasma response to OGTT particularly in vitamin B family members, mitochondrial energy metabolism, and purine metabolism. Lastly, we identified a set of metabolites significantly associated with CKD status and inflammatory markers. Together, our findings suggest CKD is associated with suppressed anabolic response to glucose challenge consistent with prior findings of impaired anabolic response to insulin in this population [52].

The application of metabolomics to the oral glucose tolerance testing demonstrated several insights into the metabolic response to a glucose challenge. First, OGTT resulted in a reduction in the concentrations of a broad range of plasma metabolites 2 hours post-glucose challenge compared to fasting suggesting an anabolic response to insulin. Second, we noted marked reduction in plasma levels of TCA cycle intermediates suggesting TCA cycle activation consistence with the known high energetic demands of protein translation and fatty acid synthesis [27] **(Table 2)**. Third, we observed a decrease in all detected amino acids and their metabolites suggesting suppression of proteolysis and cataplerosis in the overall cohort (**Table 2**). In particular, we detected significant decreases in succinate, oxaloacetate, aconitate, GDP, malonate, and ADP consistent with the known action of insulin in stimulating glucose metabolism and inhibition of catabolic pathways such as lipolysis, proteolysis and ketogenesis (**Table 2**).

We observed significant heterogeneity by CKD status in the plasma metabolome response to glucose load revealing underlying broad disruption in energy metabolism. In animal models, CKD leads to a post-insulin receptor defect contributing to IR [28, 29]. Studies have shown that IR is a major contributor to altered metabolism in CKD with adverse consequences for lipid [30], carbohydrate [31], and protein metabolism [32] impacting global energy metabolism [20]. Our investigation identified CKD-associated disruptions in GDP, ADP, and succinate; all metabolites involved in the TCA cycle. We showed these metabolites were significantly higher in CKD compared to controls post-OGTT suggesting an impaired insulin response to activate the TCA cycle (**Table 3**). These findings are also consistent with previous study demonstrating diminished TCA cycle activity in patients with non-diabetic CKD [33]. Our findings underscore the importance of IR associated with metabolic defects in CKD and its potential contribution to mitochondrial dysfunction. This association between IR and mitochondrial energy metabolism was demonstrated in a recent study suggesting that muscle specific insulin receptor knockout in a murine model impaired mitochondrial respiration, decreased ATP production, and increased reactive oxygen species [34]. This may be driven by decreased complex 1 (NADH oxidase) driven mitochondrial respiration and supercomplex assembly which may subsequently manifest as an increase in NADH/NAD ratio.

One prominent feature of mitochondrial dysfunction we noted in CKD versus control was elevated lactate-to-pyruvate (L:P) ratio. The L:P ratio is known to be in near equilibrium with the NADH/NAD ratio with an elevated lactate-to-pyruvate (L:P) ratio serving as an indicator for impaired mitochondrial function, TCA cycle, and pyruvate decarboxylase activity; a highly regulated subunit of pyruvate dehydrogenase (PDH) [35–37]. A L:P ratio of >25 has been suggestive of respiratory chain dysfunction [38]. Increased levels of pyruvate, lactate, and L:P ratio has also been shown in patients with acute kidney failure [39]. Glycolysis products, lactate and pyruvate, were increased in both groups in response to the glucose challenge signaling glycolysis activation. Lactate and pyruvate however were also both more elevated in CKD compared to the controls suggesting reduced activity of pyruvate dehydrogenase complex (PDH) (**Supplemental Table 2**). PDH is a key insulin-mediated enzyme complex with activity in the muscle which is acutely stimulated by insulin [40]. Indeed, reduced activity of insulin-stimulated PDH has been shown in skeletal muscle of patients with type 2 diabetes [40, 41].

Reduced muscle PDH activity in patients with stage 4 and 5 CKD has also been reported as a contributing factor to muscle weakness [42]. We also observed an elevated (L:P>30) lactate: pyruvate ratio in CKD compared to controls both at fasting and post-OGTT suggesting a disorder in mitochondrial respiratory chain complex (**Figure 2A)**. Taken together our findings indicate persons with CKD have impaired mitochondrial metabolism evidenced by metabolic profiles suggesting blunted activation of TCA cycle, redox imbalance, as well as disruption in the mitochondrial respiratory chain complex.

CKD was associated with disruption in purine and pyrimidine metabolism, a crucial source of necessary energy and cofactors needed for bioenergetics and biomolecular demands of metabolism in addition to serving as building blocks of DNA and RNA [43]. Purines are found in biomolecules such as ATP, GTP, cyclic AMP, NADH, and coenzyme A [44]. We noted several indicators of impairment in the purine nucleotide cycle. First, four metabolites involved in purine nucleotide cycle GDP, ADP, adenylosuccinate, and IMP all had attenuated reduction in response to the glucose challenge in CKD compared to controls suggesting an impairment in purine nucleotide cycle **(Table 3)**. Fluctuations in ATP, ADP, and AMP and corresponding changes in GTP and GDP are regulated by purine nucleotide cycle, particularly in skeletal muscle [45, 46]. During muscle contraction in the skeletal muscle a high ATP:ADP and GTP:GDP ratio is maintained during high ATP usage [34]. The purine cycle also enhances glycolysis and the TCA cycle via enhancing the rate of glycolysis by activating phosphofructokinase (PFK) and production of fumarate serving as an anaplerotic substrate supporting the TCA cycle [47, 48]. Second, to assess disruptions in its regulatory role of energy molecules during glycolytic changes, we investigated changes in GTP:GDP ratios at fasting and during OGTT (where GTP:GDP ratio is expected to increase [26, 49]) in both groups. We found that patients with CKD had similar fasting GTP:GDP ratios compared to controls, but significantly lower GTP:GDP ratio post-OGTT suggesting an impairment in the purine nucleotide cycle interfering with energy generation. Purine nucleotide cycle has not been previously explored in kidney disease and warrants further investigation in patients living with CKD given its role in various metabolic pathways fundamental in skeletal muscle energy metabolism.

CKD was also associated with disruption of metabolites in the vitamin B family with glucose challenge. Vitamin B family members are essential for metabolism involved in major metabolic pathways acting as cofactors in catabolic and anabolic pathways[50]. We found a decrease in vitamin B family derivatives in repones to OGTT in both groups. Among vitamin B family members, the most significant decrease post OGTT was observed in nicotinamide (a form of vitamin B3), important in various oxidation/reduction reactions. Compared to controls where nicotinamide levels profoundly decreased in response to glucose challenge, the levels of nicotinamide in participants with CKD remained elevated (**Table 3**) consistent with our prior findings using insulin clamp testing [51]. Our data confirms findings from these other studies showing nicotinamide and other NAD^+^ catabolites accumulating in uremic patients suggesting an insulin mediated defect to use nicotinamide during anabolic reactions. Dynamic changes in other B vitamins were also detected post glucose challenge (**Supplemental Table 2**). Biotin (vitamin B7) needed for energy metabolism, fat synthesis, amino acid metabolism and glycogen synthesis [38] significantly dropped in the controls whereas a slight increase was observed in CKD post glucose challenge (**Table 3**). Similarly, pyroxidal phosphate, the active form of vitamin B6 needed for glycine and serine metabolism in humans [38]; pantothenic acid (vitamin B5) needed for carbohydrate, protein, and fat metabolism also remained elevated post glucose challenge in CKD compared to controls [52] (**Supplemental Table 2**). The relevance of these changes is underscored by the essential role these metabolites play in metabolic pathways influenced by insulin action, such as amino acid metabolism (Vitamin B6), TCA cycle (Vitamin B5), and various redox reactions (Vitamin B3). Together, elevated levels of vitamin B family members points toward an impairment in activating insulin mediated anabolic pathways involving accumulation of these cofactors in patients with CKD.

There were several metabolic profile changes in response to the glucose challenge characteristic of CKD [53] and levels of CKD associated inflammation markers identified using WGCNA analysis [54, 55]. First, a module consisting of malonate, leucic acid, IMP, kynurenate, uric acid and inositol had a significant positive correlation with plasma creatinine, plasma cystatin C, CKD status, and a significant negative correlation with eGFR (**Figure 3B**). This module also had a significant positive correlation with TNF-α levels (**Figure 3C**). Interestingly, uremic retention solutes and uremic toxins including inositol, malonate, and uric acid make up a considerable number of correlating metabolites in this module.

Inositol is a biologically relevant metabolite involved in energy metabolism and considered a uremic retention solute. It was attenuated post-OGTT in the CKD group compared to controls, and it’s also part of the green module correlating with CKD status and TNF-α levels. Inositol is a polyol produced from glucose-6-phoshate and a water-soluble low molecular weight solute playing an important role in a cellular signaling [56]. In humans, the kidney is the main organ responsible for biosynthesis and catabolism of myo-inositol [57, 58]. Serum inositol levels in as been negatively correlation with GFR [59–61] and implicated to have an adverse impact on renal progression. Among patients with type 2 diabetes, elevated plasma concentration of myo-inositol has been associated with greater likelihood of progression to end stage renal disease (ESRD) [62]. An *in vitro* study has also shown inositol as an inhibitor of mitochondrial fission by direct inactivation of 5’-AMP-activated protein kinase (AMPK) [63], a primary energy sensor and regulator of cellular metabolism [64]. Our study, confirms and builds on the previous studies confirming elevated myo-inositol levels in patients with non-diabetic CKD. Further studies are needed to investigate the role of altered inositol metabolism in defective mitochondrial dynamics contributing to mitochondrial dysfunction and cell damage in CKD.

Two other known uremic toxins characterizing differences in the response to glucose challenge in CKD compared to controls were malonate and uric acid. Elevated malonate, a potent inhibitor of cellular respiration, has been reported in ESRD patients[65, 66]. Malonate binds to the active site of succinate dehydrogenase (Complex II) of the electron transport chain without reacting thus competing with the usual substrate succinate and disrupting oxidative phosphorylation contributing to mitochondrial dysfunction in CKD [67]. High serum uric acid levels are very common in CKD patients [68] and has been shown to be correlated with greater IR, greater C-reactive protein levels, and lower eGFR in patients with CKD [69, 70]. Uric acid activates NADPH oxidase enzyme increasing intracellular oxidative stress, mitochondrial injury, and ATP depletion [71, 72]. The protein bound solute kynurenate was also significantly altered post glucose challenge in the green module (**Table 3, Figure 3B, Figure 3C**). Elevated serum kynurenate levels have been associated with reduced kidney function[73], greater TNF-α levels [74] and greater IR [75] and is a well-documented marker for kidney disease [73, 74, 76, 77]. In summary, WGCNA analysis suggests uremic toxins are strongly associated with disruption in response to glucose challenge in CKD compared to controls and linked to inflammatory markers in our cohort.

Our study has several notable strengths but also limitations. First, we assessed dynamic changes in response to glucose challenge and accounted for several potential confounding factors by adjusting for age, sex, race, body weight, and batch in our analysis. Second, we applied targeted metabolic profiling to assess a broad range of attenuated metabolic pathways impacted by physiological effects of insulin and identified its potential disturbed mechanisms in CKD. Our study was not without limitations. We relied on plasma metabolite levels, so we had no direct evidence of metabolic alterations inside the cells or any specific tissue. Differences in gut microbiome can influence circulating metabolites and the changes we observed upon glucose loading could be in part due to differences in intestinal microbiome. We can’t rule out the potential impact of residual confounding by differences in unmeasured characteristics between CKD and controls. Finally, we can’t precisely identify the impact of potential differences in the incretin induced secretion of insulin between our two groups. However, our data confirms prior published study from our cohort where we demonstrated no significant difference in the plasma insulin levels and insulin sensitivity during OGTT comparing CKD to controls (**Figure 3A and 3B**) [22]. Further studies are necessary to interrogate the association of reduced activation of TCA cycle, redox imbalance, and electron transport chain efficiency in human tissues especially in the skeletal muscle.

In summary, our results show that numerous plasma metabolites are altered in response to oral glucose challenge in CKD. We showed that response to glucose challenge in CKD is associated with disruption in plasma levels of TCA cycle intermediates, purine nucleotide cycle metabolites, and vitamin B family members relative to controls. This abnormal metabolic profile in response to glucose challenge adds to prior studies indicating the IR in CKD is predominantly characterized by a depressed anabolic response and disruption in energy metabolism. We also showed that disruptions in plasma metabolic profile in response to glucose load, in particular uremic toxins and retention solutes, are the main metabolites correlating with CKD signature and inflammatory biomarkers. Given recent evidence for clinical benefits of insulin sensitizers, there is an urgent need for future studies interrogating the pleotropic effects of insulin sensitizers on mitochondrial energy metabolism and anabolism in CKD.

## Methods

### Population

The Study of Glucose and Insulin in Renal Disease (SUGAR) is a cross-sectional study of glucose and insulin metabolism in moderate-to-severe nondiabetic CKD. From 2011– 2014, we recruited 98 participants from Nephrology and Primary Care clinics associated with the University of Washington and neighboring institutions in Seattle (Washington, USA). Among the 98 recruited participants, 95 had adequate plasma samples collected for metabolomics, and of these, 61 had plasma samples before and after oral glucose tolerance testing(41 CKD and 20 controls) [22]. Sixty-one participants were included from the SUGAR study where 41 of them were moderate to severe non-diabetic CKD patients (eGFR < 60 ml/min per 1.73 m^2^) and 20 healthy controls (eGFR > 60 ml/min per 1.73 m^2^). Plasma lipid and non-lipid metabolites were measured after overnight fasting, 2h after a 75-gram oral glucose load for OGTT and during the clamp as reported previously [52]. Plasma biomarkers of kidney function and inflammation were measured in the fasting blood. Albumin excretion rate was calculated from 24h urine sample. A detailed description of the study design and distribution of the clinical phenotypes between CKD patients and healthy controls has been reported earlier [22].

### Metabolomics

**Targeted metabolomics based in a** liquid chromatography-tandem mass spectroscopy (LC-MS/MS) platform was performed at the Northwest Metabolomics Research Center as described previously [78]. The LC-MS/MS analyses were performed using an Agilent 1260 LC (Agilent Technologies) with a SeQuant ZIC-cHILIC column (150 × 2.1 mm × particle size 3.0 μm, Merck KGaA), which was coupled to a AB Sciex QTrap 5500 MS (AB Sciex) system equipped with a standard electro-spray ionization (ESI) source. The details for sample preparation, sample collection times and reagents has been described earlier [51].

### Statistical analysis

Plasma non-lipid metabolite data were available from 61 subjects at fasting and 2h post-oral glucose load. Raw metabolite data were converted to ratio of 2h post-oral glucose load and corresponding fasting values and checked for batch effect of the ratio values. All clinical and metabolite data were checked for normality. To account for drift of sample preparation batches, raw metabolite data was normalized using Systematic Error Removal Using Random Forest (SERRF)[79]. To account for the correlation of measurements within participants, we examined the fold changes associated with the OGTT procedure via a linear mixed model with random intercepts, regressing the log transformed SERRF normalized metabolite on the sample type (during OGTT versus fasting sample), adjusting for age, sex, race/ethnicity, body weight and batch. To evaluate whether the effect of the oral glucose challenge procedure differed between CKD and non-CKD participants, we used linear mixed effect modeling with random intercepts where SERRF normalized metabolites were regressed on a sample type (OGTT vs fasting), CKD status, and their interaction, additionally adjusting for covariates listed above. Data are presented as means ± SD unless otherwise indicated. Statistical analysis was performed using R 3.6.1 for windows release [80]. P-values were adjusted for multiple comparisons using the “Benjamini Hochberg” appraoch, and an adjusted p-value <0.05 was considered significant for all analyses unless stated otherwise.

Pathway-associated metabolite sets enrichment and metabolite pathway analysis were performed using MetaboAnalystR v4.0 [81] with the KEGG human metabolite database [82]. Pathway analysis using SERRF normalized pre OGTT and post OGTT metabolite level was used to evaluate metabolic changes in response to OGTT in the entire cohort. Significantly altered metabolic pathway by CKD status was determined by comparing the changes in metabolite levels calculated by subtracting log-transformed post OGTT SERRF normalized values to pre OGTT time points.

Differences in insulin sensitivity (Matsuda index) and plasma metabolites including GTP:GDP ratio and lactate: pyruvate ratio between fasting and 2h post-oral glucose load were evaluated separately by Mann-Whitney U test using GraphPad Prism 9.0 (GraphPad Software, Inc., San Diego, CA).

Metabolite data were checked for excessive missing values by using the R package WGCNA’s [83] “goodSampleGenes” test. The correlation between kidney biomarkers was determined by Spearman correlation. The ratio of plasma metabolites post oral glucose to pre glucose challenge was used for correlation network analysis using WGCNA R package. A soft threshold approach was used with a power of 6 (based on scales free topology) in a WGCNA default unsigned network with dynamic tree cutting (deep split = 2) and a min Module Size = 3 as parameters for the dynamic tree cut function [84]. The module eigengene, defined as the first principal component (PC) of a module’s metabolite concentration matrix, was used to calculate the Spearman correlation between a metabolite module and kidney biomarkers.

## Author Contributions

Conceptualization was contributed by AA, BR, MNH, LRZ, DR, and IHDB. Methodology was contributed by AA, MNH, LRZ, DR, and BR. Formal analysis was contributed by AA and MNH. Investigation was contributed by BR, MNH, and DR. Resources were contributed by IHDB, BR, and DR. Data curation was contributed by AA and MNH. Writing the original draft was contributed by AA, BR, and MNH. Writing the review and editing were contributed by AA, BR, MNH, LRZ, IHDB, BJB, JG, LRD, MC, DR. Visualization was contributed by AA and MNH. Supervision was contributed by IHDB, BR, BJB, and DR. Project administration was contributed by IHDB and DR. Funding acquisition was contributed by BR, IHDB, DR, and BJB.

## Data Availability

All data produced in the present study are available upon reasonable request to the authors

## Acknowledgements

This project was funded in part by an unrestricted grant from the Northwest Kidney Centers and R01DK087726, R01DK087726-S1, R01DK129793-01 (BR), R01DK087726, R01DK087726- S1, K01 DK102851 (JAA, IHDB), K23DK100533 (JLG), K24 DK096574 (TRZ), and P30 DK017047 (University of Washington Diabetes Research Center) and Dialysis Clinics Incorporated, C-4122 (BR).

## Supplemental Figures

**Supplemental table 1.**
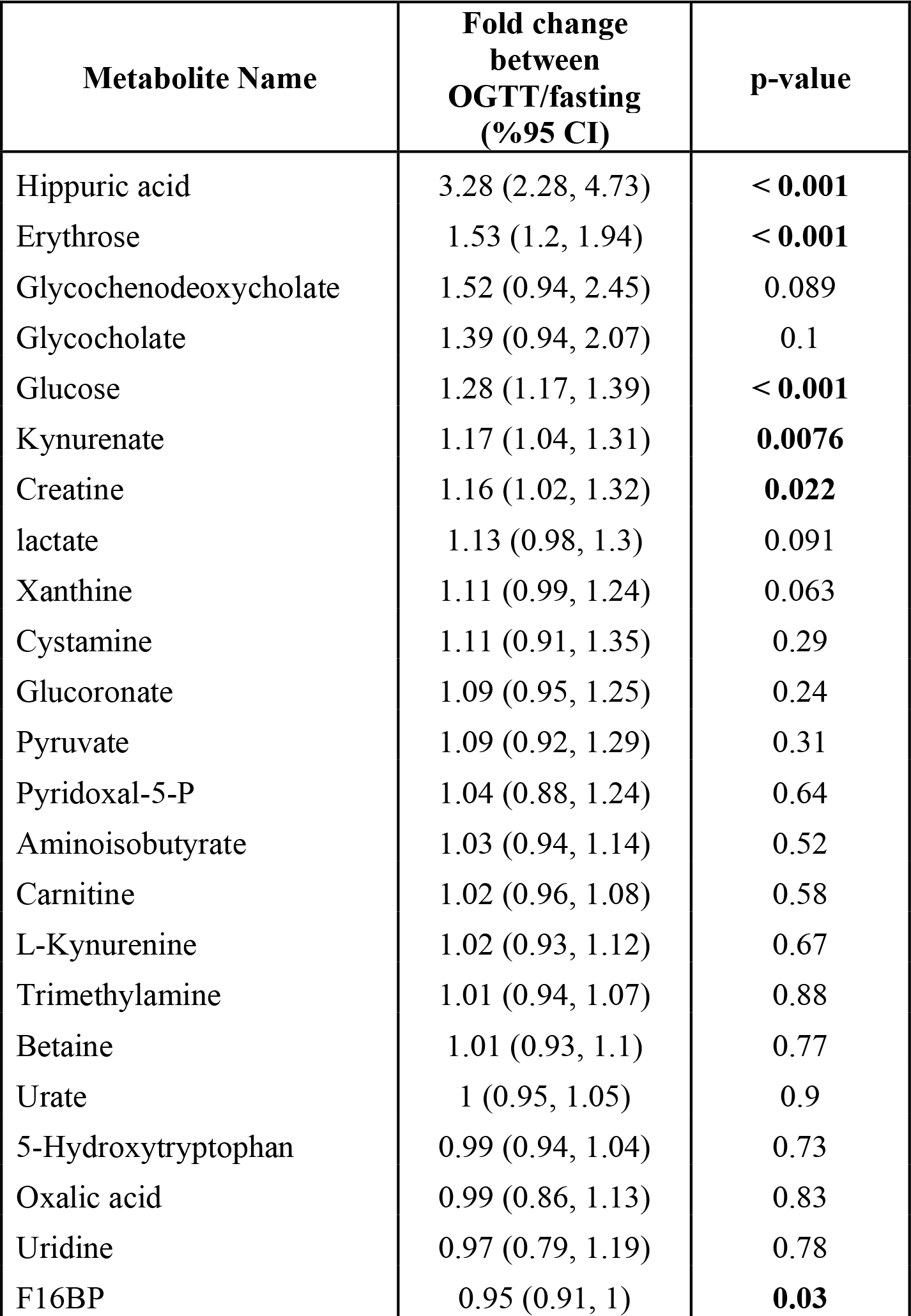
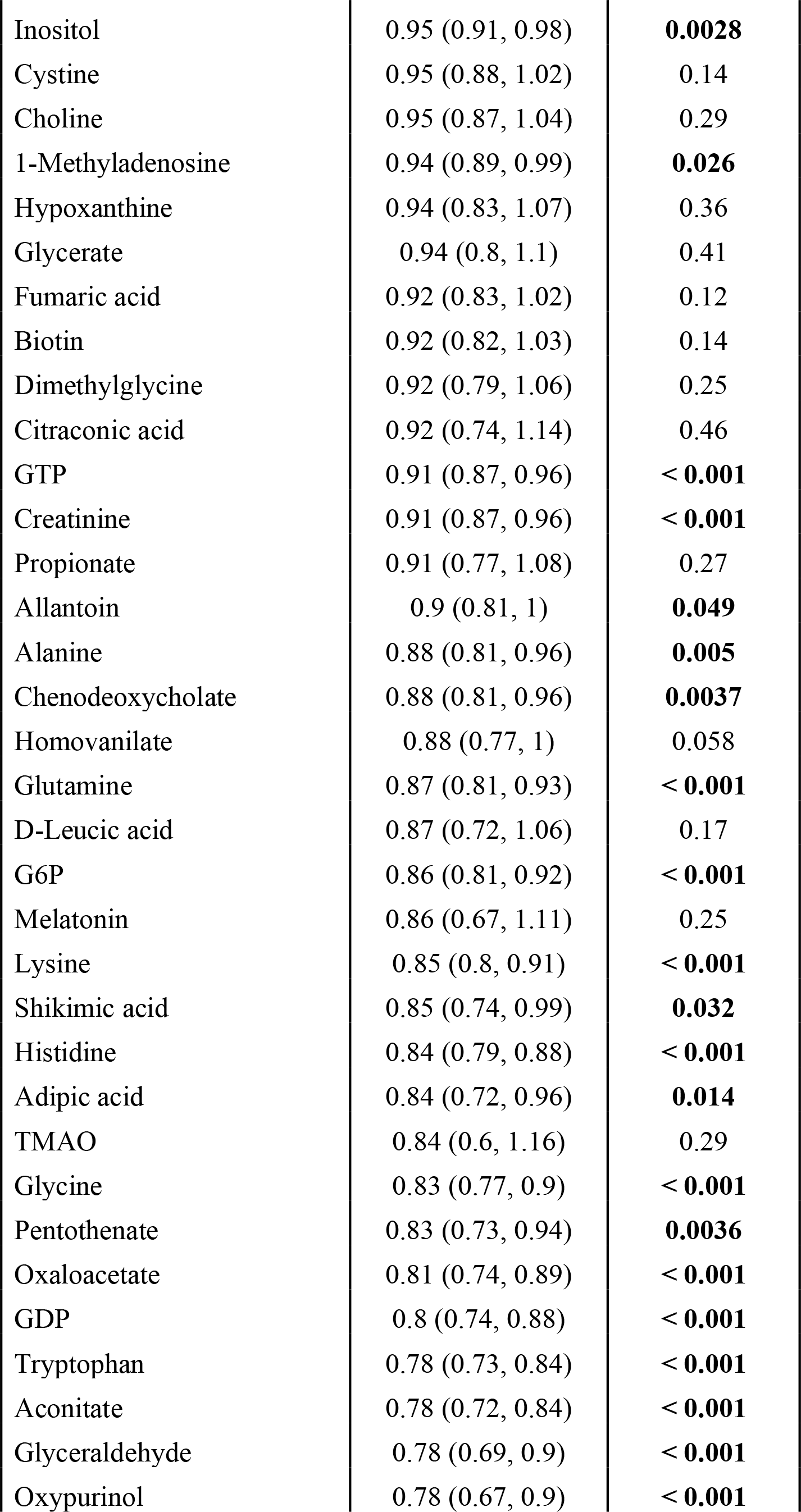
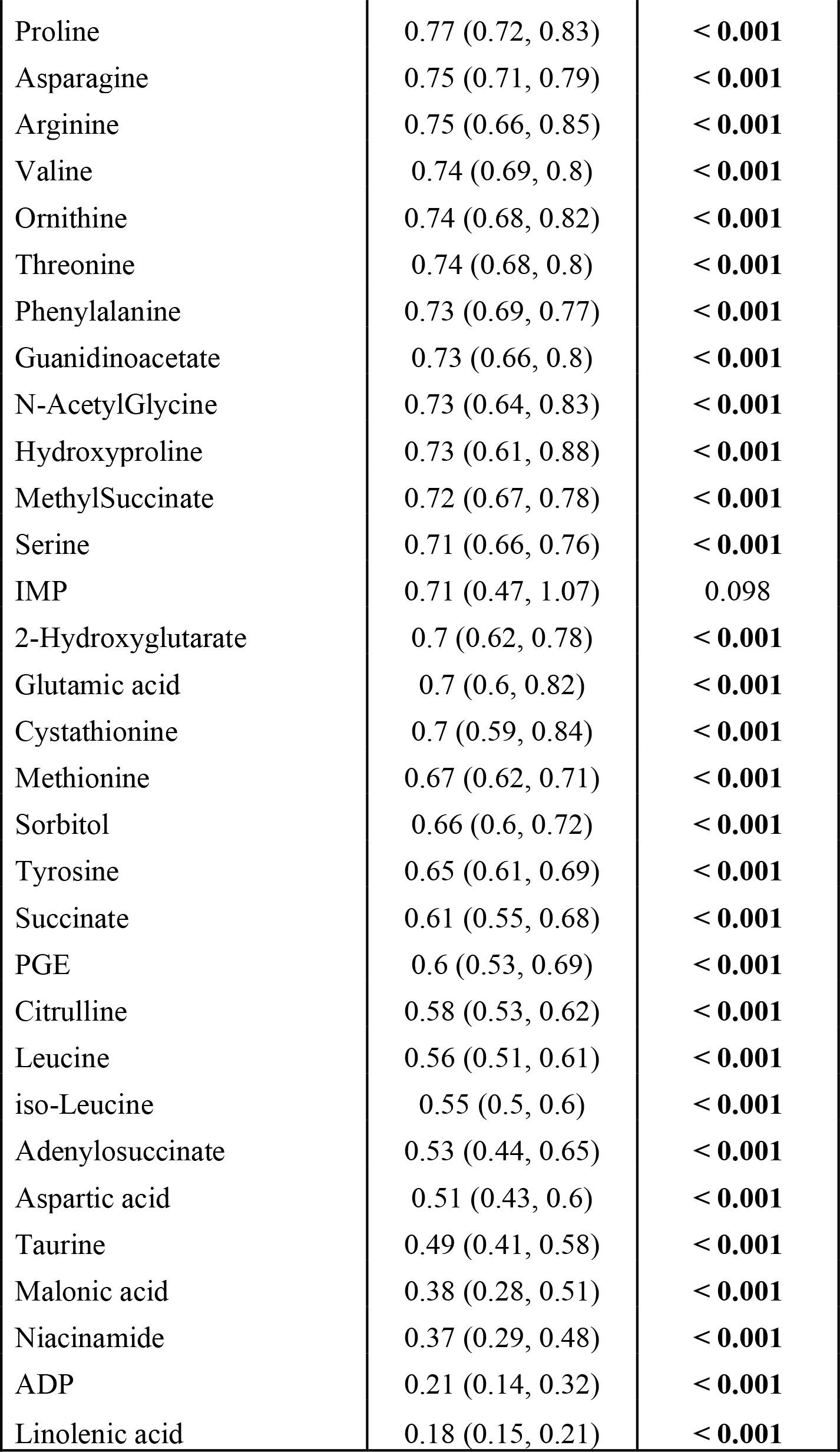
Differences in plasma metabolites in response to glucose load compared to fasting. Results are from a regression SERRF normalized metabolites on sample type (OGTT vs fasting) adjusted for age, sex, race, weight, and batch. A p-value of <0.05 was used to determine significance (in bold). The fold change is in response to oral glucose is the adjusted fold change associated with OGTT compared to fasting state (e.g., a fold change of 1.28 indicates a 28% increase in metabolite levels).

**Supplemental table 2.**
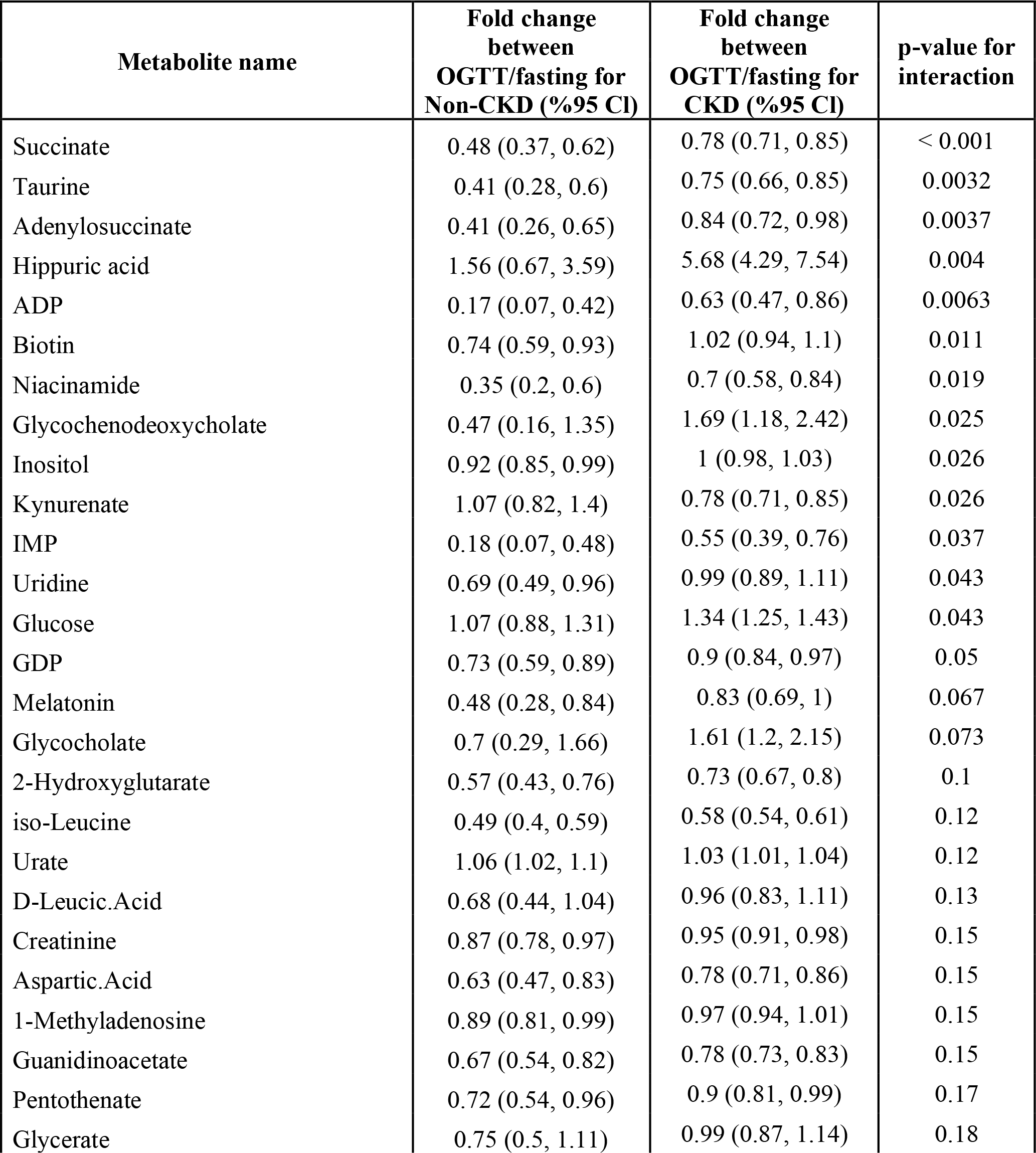
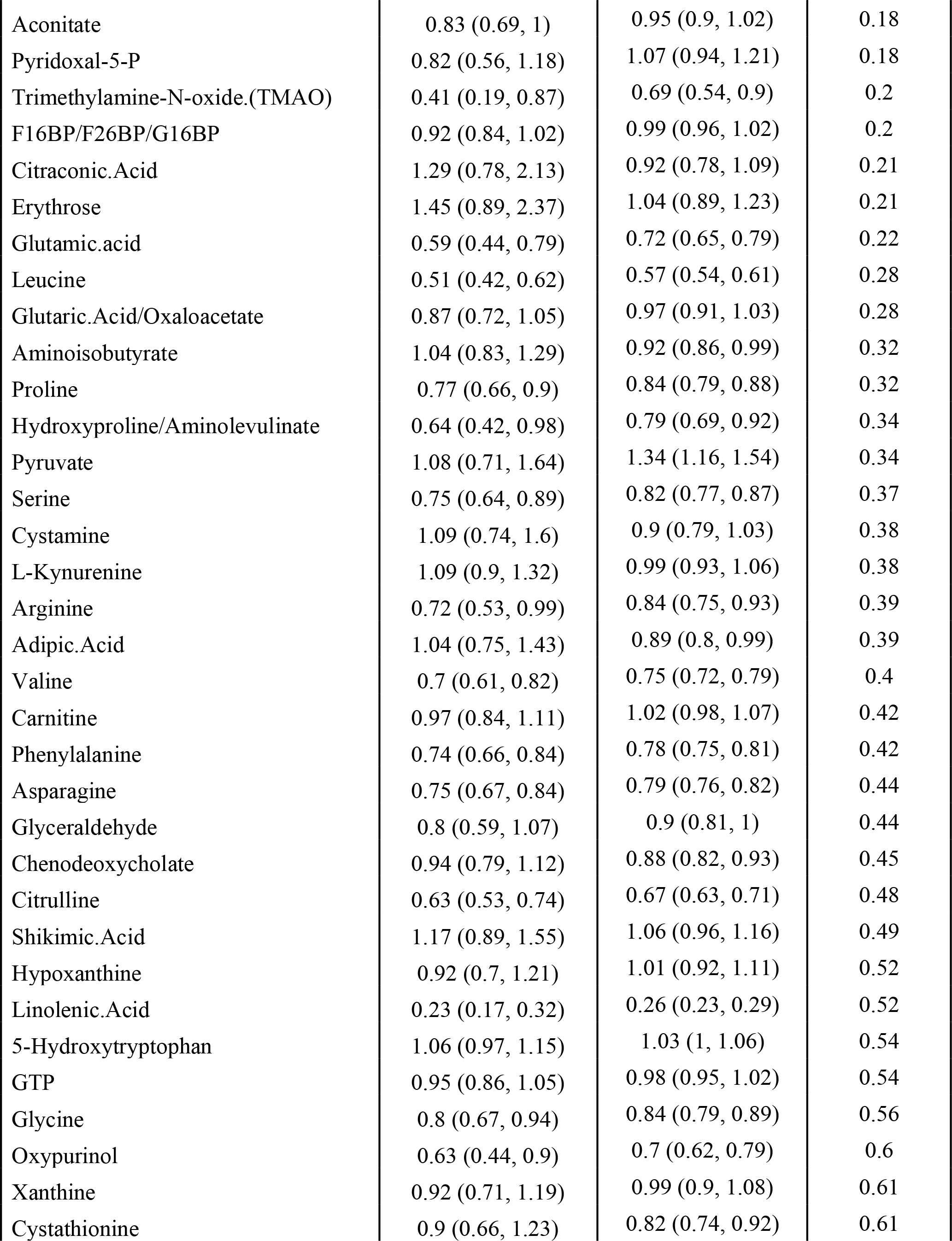
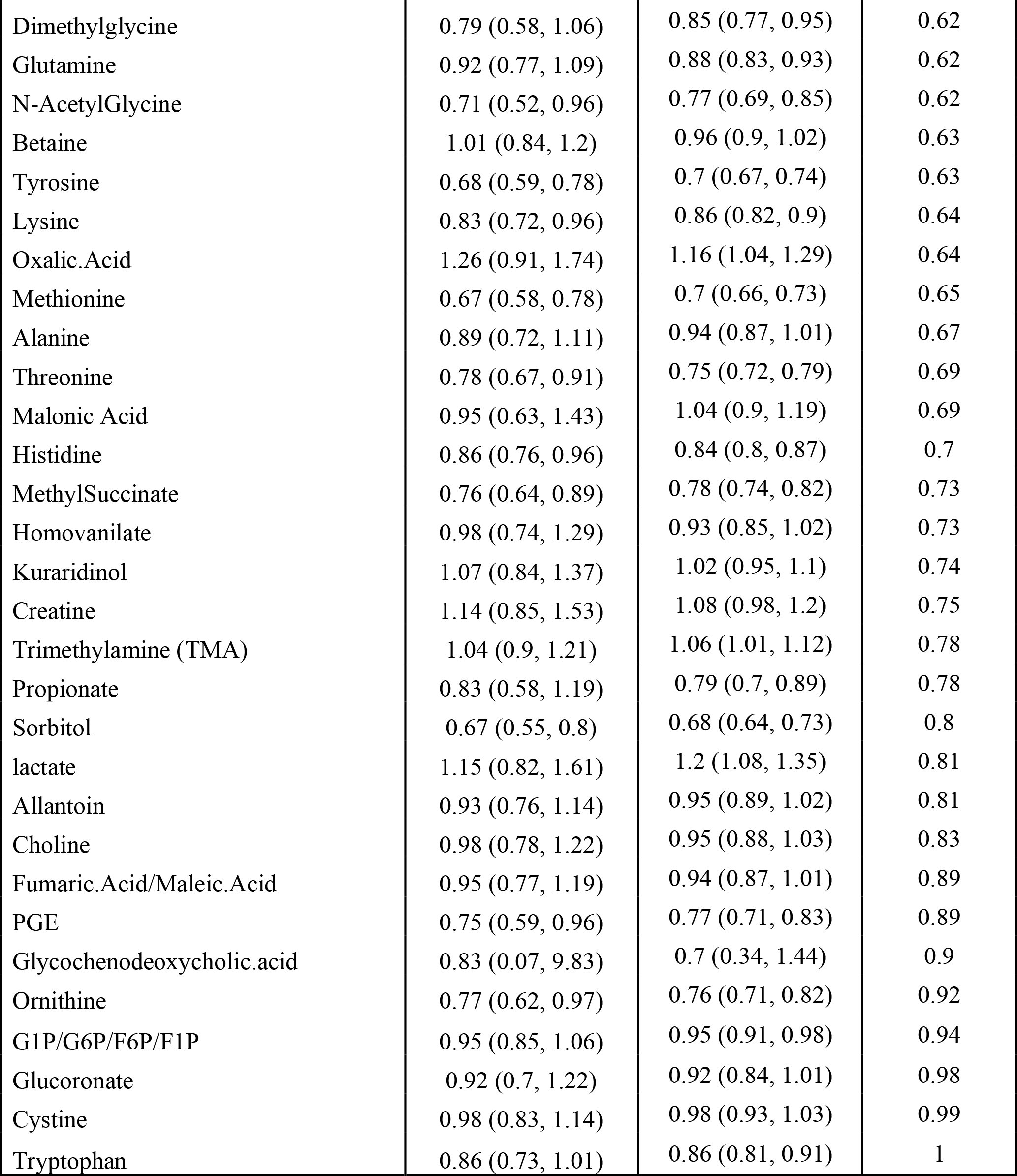
Differences in plasma metabolic response post glucose challenge by CKD status. Result from regression analysis using SERFF normalized on sample type (fasting vs OGTT) by CKD status adjusted for age, sex, race, weight, and batch. Fold changes represent changes in metabolite levels with fasting levels after glucose load. (e.g. a fold change of 0.92 indicates a reduction of %8 in metabolite level. P-value for interaction represents the heterogeneity in fold change by disease status.

**Supplemental table 3.**
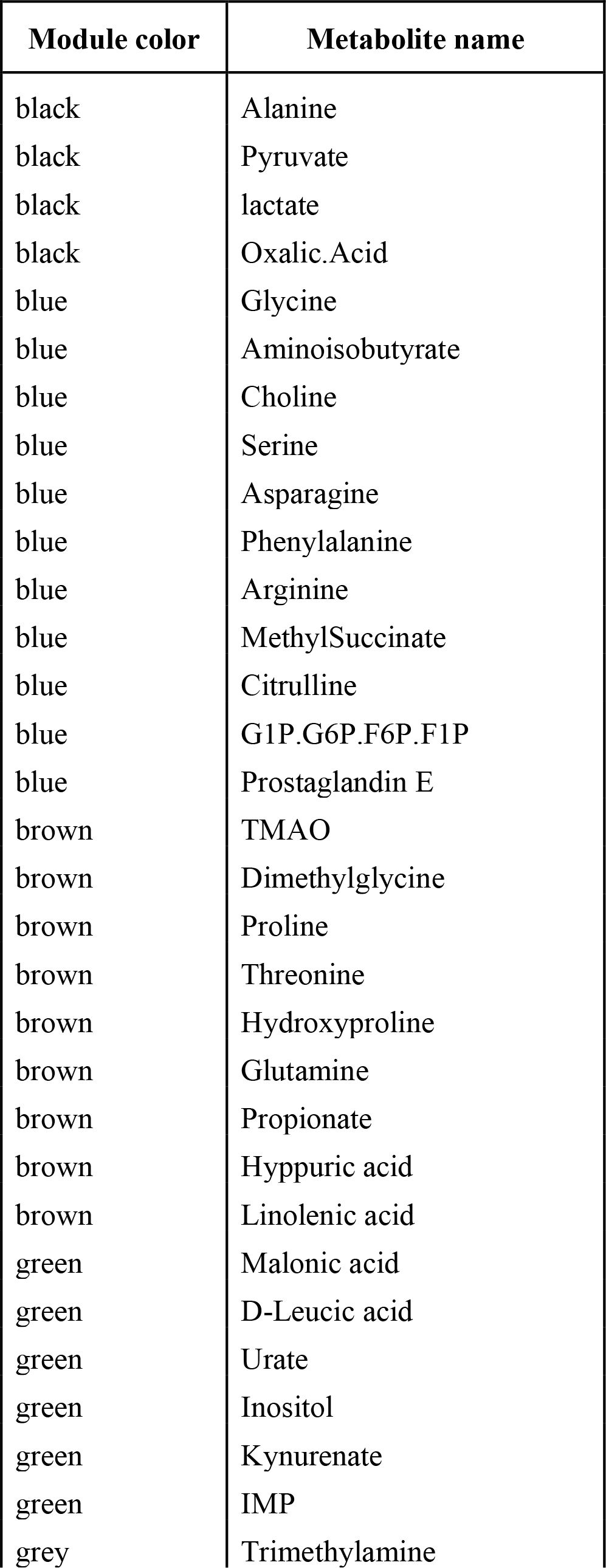
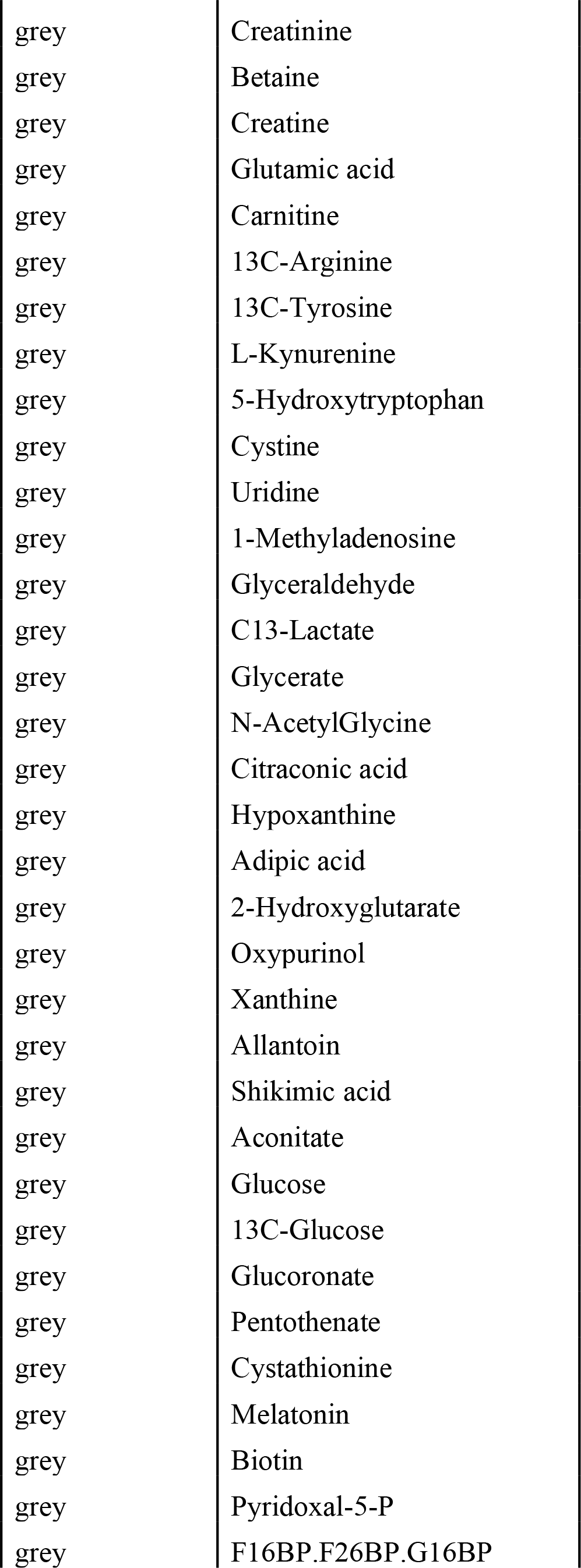
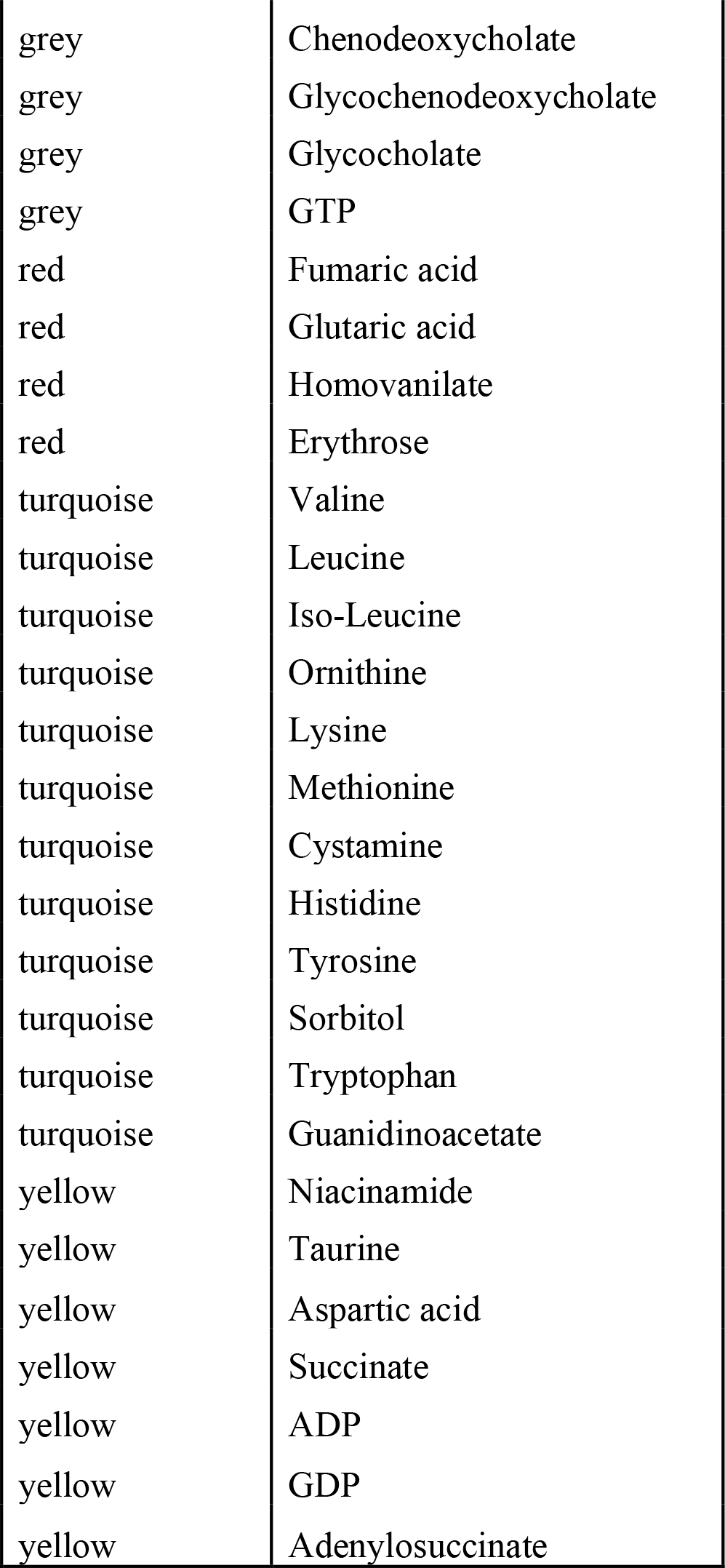
The list of metabolites in each module from the WGCNA analysis.

## References

1. Bikbov, B., et al., Global, regional, and national burden of chronic kidney disease, 1990&#x2013;2017: a systematic analysis for the Global Burden of Disease Study 2017. The Lancet, 2020. 395(10225): p. 709–733.

2. Spoto, B., A. Pisano, and C. Zoccali, Insulin resistance in chronic kidney disease: a systematic review. Am J Physiol Renal Physiol, 2016. 311(6): p. F1087–f1108.

3. de Luca, C. and J.M. Olefsky, Inflammation and insulin resistance. FEBS letters, 2008. 582(1): p. 97–105.

4. Hurrle, S. and W.H. Hsu, The etiology of oxidative stress in insulin resistance. Biomedical journal, 2017. 40(5): p. 257–262.

5. Is fasting glucose sufficient to define diabetes? Epidemiological data from 20 European studies. The DECODE-study group. European Diabetes Epidemiology Group. Diabetes Epidemiology: Collaborative analysis of Diagnostic Criteria in Europe. Diabetologia, 1999. 42(6): p. 647–54.

6. de Vegt, F., et al., Hyperglycaemia is associated with all-cause and cardiovascular mortality in the Hoorn population: the Hoorn Study. Diabetologia, 1999. 42(8): p. 926–31.

7. Henriksen, E.J., M.K. Diamond-Stanic, and E.M. Marchionne, Oxidative stress and the etiology of insulin resistance and type 2 diabetes. Free Radic Biol Med, 2011. 51(5): p. 993–9.

8. Kanety, H., et al., Tumor necrosis factor alpha-induced phosphorylation of insulin receptor substrate-1 (IRS-1). Possible mechanism for suppression of insulin-stimulated tyrosine phosphorylation of IRS-1. J Biol Chem, 1995. 270(40): p. 23780–4.

9. Senn, J.J., et al., Interleukin-6 induces cellular insulin resistance in hepatocytes. Diabetes, 2002. 51(12): p. 3391–9.

10. Liao, M.T., et al., Insulin resistance in patients with chronic kidney disease. J Biomed Biotechnol, 2012. 2012: p. 691369.

11. Koppe, L., et al., p-Cresyl sulfate promotes insulin resistance associated with CKD. J Am Soc Nephrol, 2013. 24(1): p. 88–99.

12. Wright, V.P., P.J. Reiser, and T.L. Clanton, Redox modulation of global phosphatase activity and protein phosphorylation in intact skeletal muscle. J Physiol, 2009. 587(Pt 23): p. 5767–81.

13. Xue, P., et al., Adipose deficiency of Nrf2 in ob/ob mice results in severe metabolic syndrome. Diabetes, 2013. 62(3): p. 845–54.

14. de Vinuesa, S.G., et al., Insulin resistance, inflammatory biomarkers, and adipokines in patients with chronic kidney disease: effects of angiotensin II blockade. J Am Soc Nephrol, 2006. 17(12 Suppl 3): p. S206-12.

15. Walker, B.G., et al., INHIBITION OF INSULIN BY ACIDOSIS. Lancet, 1963. 2(7315): p. 9641-5.

16. Thomas, S.S., L. Zhang, and W.E. Mitch, Molecular mechanisms of insulin resistance in chronic kidney disease. Kidney International, 2015. 88(6): p. 1233–1239.

17. Shi, J., et al., Cytokines and Abnormal Glucose and Lipid Metabolism. Frontiers in Endocrinology, 2019. 10(703).

18. Slee, A.D., Exploring metabolic dysfunction in chronic kidney disease. Nutrition & Metabolism, 2012. 9(1): p. 36.

19. Kestenbaum, B., et al., Impaired skeletal muscle mitochondrial bioenergetics and physical performance in chronic kidney disease. JCI Insight, 2020. 5(5).

20. Thomas, S.S., L. Zhang, and W.E. Mitch, Molecular mechanisms of insulin resistance in chronic kidney disease. Kidney Int, 2015. 88(6): p. 1233–1239.

21. Rahhal, M.-N., et al., Disturbances in Insulin–Glucose Metabolism in Patients With Advanced Renal Disease With and Without Diabetes. The Journal of Clinical Endocrinology & Metabolism, 2019. 104(11): p. 4949–4966.

22. De Boer, I.H., et al., Impaired glucose and insulin homeostasis in moderate-severe CKD. Journal of the American Society of Nephrology, 2016. 27(9): p. 2861–2871.

23. Reaven, G.M., et al., Insulin resistance and insulin secretion are determinants of oral glucose tolerance in normal individuals. Diabetes, 1993. 42(9): p. 1324–32.

24. Perley, M.J. and D.M. Kipnis, Plasma insulin responses to oral and intravenous glucose: studies in normal and diabetic sujbjects. J Clin Invest, 1967. 46(12): p. 1954–62.

25. McIntyre, N., C.D. Holdsworth, and D.S. Turner, NEW INTERPRETATION OF ORAL GLUCOSE TOLERANCE. Lancet, 1964. 2(7349): p. 20-1.

26. Detimary, P., G. Van den Berghe, and J.C. Henquin, Concentration dependence and time course of the effects of glucose on adenine and guanine nucleotides in mouse pancreatic islets. J Biol Chem, 1996. 271(34): p. 20559–65.

27. Golshani-Hebroni, S.G. and S.P. Bessman, Hexokinase binding to mitochondria: a basis for proliferative energy metabolism. J Bioenerg Biomembr, 1997. 29(4): p. 331–8.

28. Bailey, J.L., et al., Chronic kidney disease causes defects in signaling through the insulin receptor substrate/phosphatidylinositol 3-kinase/Akt pathway: implications for muscle atrophy. J Am Soc Nephrol, 2006. 17(5): p. 1388–94.

29. Smith, D. and R.A. DeFronzo, Insulin resistance in uremia mediated by postbinding defects. Kidney Int, 1982. 22(1): p. 54–62.

30. Tangvarasittichai, S., Oxidative stress, insulin resistance, dyslipidemia and type 2 diabetes mellitus. World J Diabetes, 2015. 6(3): p. 456–80.

31. Ikee, R., et al., Glucose metabolism, insulin resistance, and renal pathology in non-diabetic chronic kidney disease. Nephron Clin Pract, 2008. 108(2): p. c163–8.

32. Siew, E.D. and T.A. Ikizler, Determinants of insulin resistance and its effects on protein metabolism in patients with advanced chronic kidney disease. Contrib Nephrol, 2008. 161: p. 138–144.

33. Hallan, S., et al., Metabolomics and Gene Expression Analysis Reveal Down-regulation of the Citric Acid (TCA) Cycle in Non-diabetic CKD Patients. EBioMedicine, 2017. 26: p. 68–77.

34. Bhardwaj, G., et al., Insulin and IGF-1 receptors regulate complex I-dependent mitochondrial bioenergetics and supercomplexes via FoxOs in muscle. J Clin Invest, 2021. 131(18).

35. Munnich, A., et al., Clinical presentations and laboratory investigations in respiratory chain deficiency. Eur J Pediatr, 1996. 155(4): p. 262–74.

36. Robinson, B.H., et al., Disorders of pyruvate carboxylase and the pyruvate dehydrogenase complex. J Inherit Metab Dis, 1996. 19(4): p. 452–62.

37. Shoffner, J.M., An introduction: oxidative phosphorylation diseases. Semin Neurol, 2001. 21(3): p. 237–50.

38. Lane, M., et al., Mitochondrial dysfunction in liver failure requiring transplantation. J Inherit Metab Dis, 2016. 39(3): p. 427–436.

39. Ando, M. and K. Shimizu, [Acute renal failure with lactic acidosis]. Nihon Jinzo Gakkai Shi, 1990. 32(6): p. 729–37.

40. Mandarino, L.J., et al., Adipocyte glycogen synthase and pyruvate dehydrogenase in obese and type II diabetic subjects. American Journal of Physiology-Endocrinology and Metabolism, 1986. 251(4): p. E489–E496.

41. Kelley, D.E., M. Mokan, and L.J. Mandarino, Intracellular Defects in Glucose Metabolism in Obese Patients With NIDDM. Diabetes, 1992. 41(6): p. 698–706.

42. Xu, C., et al., Reduction of mitochondria and up regulation of pyruvate dehydrogenase kinase 4 of skeletal muscle in patients with chronic kidney disease. Nephrology (Carlton), 2020. 25(3): p. 230–238.

43. Pedley, A.M. and S.J. Benkovic, A New View into the Regulation of Purine Metabolism: The Purinosome. Trends in Biochemical Sciences, 2017. 42(2): p. 141–154.

44. Kumari, A., Chapter 17 - Purine Structures, in Sweet Biochemistry, A. Kumari, Editor. 2018, Academic Press. p. 89–91.

45. Tornheim, K., Oscillations of the glycolytic pathway and the purine nucleotide cycle. J Theor Biol, 1979. 79(4): p. 491–541.

46. Tornheim, K. and J.M. Lowenstein, The purine nucleotide cycle. Control of phosphofructokinase and glycolytic oscillations in muscle extracts. J Biol Chem, 1975. 250(16): p. 6304–14.

47. Arinze, I.J., Facilitating understanding of the purine nucleotide cycle and the one-carbon pool: Part I: The purine nucleotide cycle. Biochemistry and Molecular Biology Education, 2005. 33(3): p. 165–168.

48. Bhagavan, N.V., CHAPTER 27 - Nucleotide Metabolism, in Medical Biochemistry (Fourth Edition), N.V. Bhagavan, Editor. 2002, Academic Press: San Diego. p. 615–644.

49. Windisch, R.M., P.R. Pax, and M.M. Bracken, Variations in blood ATP after oral administration of glucose, in individuals diagnosed as normal, equivocal, or diabetic according to the glucose tolerance sum principle. Clin Chem, 1970. 16(11): p. 941–4.

50. Vitamins Important for Metabolism. 2020.

51. Roshanravan, B., et al., Chronic kidney disease attenuates the plasma metabolome response to insulin. JCI Insight, 2018. 3(16).

52. Institute of Medicine Standing Committee on the Scientific Evaluation of Dietary Reference, I., O.B.V. its Panel on Folate, and Choline, *The National Academies Collection: Reports funded by National Institutes of Health*, in *Dietary Reference Intakes for Thiamin, Riboflavin, Niacin, Vitamin B(6), Folate, Vitamin B(12), Pantothenic Acid, Biotin, and Choline*. 1998, National Academies Press (US) Copyright © 1998, National Academy of Sciences.: Washington (DC).

53. Lopez-Giacoman, S. and M. Madero, Biomarkers in chronic kidney disease, from kidney function to kidney damage. World J Nephrol, 2015. 4(1): p. 57–73.

54. Lavín-Gómez, B.A., et al., Inflammation markers, chronic kidney disease, and renal replacement therapy. Adv Perit Dial, 2011. 27: p. 33–7.

55. Muslimovic, A., et al., Inflammatory Markers and Procoagulants in Chronic Renal Disease Stages 1-4. Med Arch, 2015. 69(5): p. 307–10.

56. Di Paolo, G. and P. De Camilli, Phosphoinositides in cell regulation and membrane dynamics. Nature, 2006. 443(7112): p. 651–7.

57. Howard, C.F., Jr. and L. Anderson, Metabolism of myo-inositol in animals. II. Complete catabolism of myo-inositol-14C by rat kidney slices. Arch Biochem Biophys, 1967. 118(2): p. 332–9.

58. Lewin, L.M., et al., Studies on the metabolic role of myo-inositol. Distribution of radioactive myo-inositol in the male rat. Biochem J, 1976. 156(2): p. 375–80.

59. Sekula, P., et al., A Metabolome-Wide Association Study of Kidney Function and Disease in the General Population. J Am Soc Nephrol, 2016. 27(4): p. 1175–88.

60. Sui, W., et al., A proton nuclear magnetic resonance-based metabonomics study of metabolic profiling in immunoglobulin a nephropathy. Clinics (Sao Paulo), 2012. 67(4): p. 363–73.

61. Qi, S., et al., A pilot metabolic profiling study in serum of patients with chronic kidney disease based on (1) H-NMR-spectroscopy. Clin Transl Sci, 2012. 5(5): p. 379–85.

62. Niewczas, M.A., et al., Uremic solutes and risk of end-stage renal disease in type 2 diabetes: metabolomic study. Kidney Int, 2014. 85(5): p. 1214–24.

63. Hsu, C.C., et al., Inositol serves as a natural inhibitor of mitochondrial fission by directly targeting AMPK. Mol Cell, 2021. 81(18): p. 3803–3819.e7.

64. Trefts, E. and R.J. Shaw, AMPK: restoring metabolic homeostasis over space and time. Mol Cell, 2021. 81(18): p. 3677–3690.

65. Baptista, A.L., et al., Potential Biomarkers of the Turnover, Mineralization, and Volume Classification: Results Using NMR Metabolomics in Hemodialysis Patients. JBMR Plus, 2020. 4(7): p. e10372.

66. Rhee, E.P., et al., Metabolite profiling identifies markers of uremia. J Am Soc Nephrol, 2010. 21(6): p. 1041–1051.

67. Pardee, A.B. and V.R. Potter, Malonate inhibition of oxidations in the Krebs tricarboxylic acid cycle. J Biol Chem, 1949. 178(1): p. 241–50.

68. Johnson, R.J., et al., Uric acid and chronic kidney disease: which is chasing which? Nephrol Dial Transplant, 2013. 28(9): p. 2221–8.

69. Kanbay, M., et al., Serum Uric Acid Level and Endothelial Dysfunction in Patients with Nondiabetic Chronic Kidney Disease. American Journal of Nephrology, 2011. 33(4): p. 298–304.

70. Sharaf El Din, U.A.A., M.M. Salem, and D.O. Abdulazim, Uric acid in the pathogenesis of metabolic, renal, and cardiovascular diseases: A review. J Adv Res, 2017. 8(5): p. 537–548.

71. Sánchez-Lozada, L.G., et al., Uric acid-induced endothelial dysfunction is associated with mitochondrial alterations and decreased intracellular ATP concentrations. Nephron Exp Nephrol, 2012. 121(3-4): p. e71–8.

72. Cristóbal-García, M., et al., Renal oxidative stress induced by long-term hyperuricemia alters mitochondrial function and maintains systemic hypertension. Oxid Med Cell Longev, 2015. 2015: p. 535686.

73. Lee, H., et al., Amino Acid Metabolites Associated with Chronic Kidney Disease: An Eight-Year Follow-Up Korean Epidemiology Study. Biomedicines, 2020. 8(7): p. 222.

74. Debnath, S., et al., Tryptophan Metabolism in Patients With Chronic Kidney Disease Secondary to Type 2 Diabetes: Relationship to Inflammatory Markers. Int J Tryptophan Res, 2017. 10: p. 1178646917694600.

75. Li, L.O., et al., Early hepatic insulin resistance in mice: a metabolomics analysis. Mol Endocrinol, 2010. 24(3): p. 657–66.

76. Grams, M.E., et al., Metabolomic Alterations Associated with Cause of CKD. Clin J Am Soc Nephrol, 2017. 12(11): p. 1787–1794.

77. Konje, V.C., et al., Tryptophan levels associate with incident cardiovascular disease in chronic kidney disease. Clinical Kidney Journal, 2020. 14(4): p. 1097–1105.

78. Zhu, J., et al., Colorectal cancer detection using targeted serum metabolic profiling. J Proteome Res, 2014. 13(9): p. 4120–30.

79. Fan, S., et al., Systematic Error Removal Using Random Forest for Normalizing Large-Scale Untargeted Lipidomics Data. Anal Chem, 2019. 91(5): p. 3590–3596.

80. R Core, T., R: A Language and Environment for Statistical Computing, R Foundation for Statistical Computing, Austria, 2019. 2019, URL http://www.R-project.org.

81. Chong, J., et al., MetaboAnalyst 4.0: towards more transparent and integrative metabolomics analysis. Nucleic Acids Res, 2018. 46(W1): p. W486–W494.

82. Ogata, H., et al., Computation with the KEGG pathway database. Biosystems, 1998. 47(1-2): p. 119–128.

83. Langfelder, P. and S. Horvath, WGCNA: an R package for weighted correlation network analysis. BMC Bioinformatics, 2008. 9: p. 559.

84. Langfelder, P., B. Zhang, and S. Horvath, Defining clusters from a hierarchical cluster tree: the Dynamic Tree Cut package for R. Bioinformatics, 2007. 24(5): p. 719–720.

