## Supplemental Data for "Chronic kidney disease is associated with attenuated plasma metabolome response to oral glucose tolerance testing"

### Supplemental Figures

**Supplemental table 1.** Differences in plasma metabolites in response to glucose load compared to fasting. Results are from a regression SERRF normalized metabolites on sample type (OGTT vs fasting) adjusted for age, sex, race, weight, and batch. A p-value of <0.05 was used to determine significance (in bold). The fold change is in response to oral glucose is the adjusted fold change associated with OGTT compared to fasting state (e.g., a fold change of 1.28 indicates a 28% increase in metabolite levels).

| Metabolite Name | Fold change between OGTT/fasting (%95 CI) | p-value |
|---|---|---|
| Hippuric acid | 3.28 (2.28, 4.73) | **< 0.001** |
| Erythrose | 1.53 (1.2, 1.94) | **< 0.001** |
| Glycochenodeoxycholate | 1.52 (0.94, 2.45) | 0.089 |
| Glycocholate | 1.39 (0.94, 2.07) | 0.1 |
| Glucose | 1.28 (1.17, 1.39) | **< 0.001** |
| Kynurenate | 1.17 (1.04, 1.31) | **0.0076** |
| Creatine | 1.16 (1.02, 1.32) | **0.022** |
| lactate | 1.13 (0.98, 1.3) | 0.091 |
| Xanthine | 1.11 (0.99, 1.24) | 0.063 |
| Cystamine | 1.11 (0.91, 1.35) | 0.29 |
| Glucoronate | 1.09 (0.95, 1.25) | 0.24 |
| Pyruvate | 1.09 (0.92, 1.29) | 0.31 |
| Pyridoxal-5-P | 1.04 (0.88, 1.24) | 0.64 |
| Aminoisobutyrate | 1.03 (0.94, 1.14) | 0.52 |
| Carnitine | 1.02 (0.96, 1.08) | 0.58 |
| L-Kynurenine | 1.02 (0.93, 1.12) | 0.67 |
| Trimethylamine | 1.01 (0.94, 1.07) | 0.88 |
| Betaine | 1.01 (0.93, 1.1) | 0.77 |
| Urate | 1 (0.95, 1.05) | 0.9 |
| 5-Hydroxytryptophan | 0.99 (0.94, 1.04) | 0.73 |
| Oxalic acid | 0.99 (0.86, 1.13) | 0.83 |
| Uridine | 0.97 (0.79, 1.19) | 0.78 |
| F16BP | 0.95 (0.91, 1) | **0.03** |

| | | |
|---|---|---|
| Inositol | 0.95 (0.91, 0.98) | **0.0028** |
| Cystine | 0.95 (0.88, 1.02) | 0.14 |
| Choline | 0.95 (0.87, 1.04) | 0.29 |
| 1-Methyladenosine | 0.94 (0.89, 0.99) | **0.026** |
| Hypoxanthine | 0.94 (0.83, 1.07) | 0.36 |
| Glycerate | 0.94 (0.8, 1.1) | 0.41 |
| Fumaric acid | 0.92 (0.83, 1.02) | 0.12 |
| Biotin | 0.92 (0.82, 1.03) | 0.14 |
| Dimethylglycine | 0.92 (0.79, 1.06) | 0.25 |
| Citraconic acid | 0.92 (0.74, 1.14) | 0.46 |
| GTP | 0.91 (0.87, 0.96) | < **0.001** |
| Creatinine | 0.91 (0.87, 0.96) | < **0.001** |
| Propionate | 0.91 (0.77, 1.08) | 0.27 |
| Allantoin | 0.9 (0.81, 1) | **0.049** |
| Alanine | 0.88 (0.81, 0.96) | **0.005** |
| Chenodeoxycholate | 0.88 (0.81, 0.96) | **0.0037** |
| Homovanilate | 0.88 (0.77, 1) | 0.058 |
| Glutamine | 0.87 (0.81, 0.93) | < **0.001** |
| D-Leucic acid | 0.87 (0.72, 1.06) | 0.17 |
| G6P | 0.86 (0.81, 0.92) | < **0.001** |
| Melatonin | 0.86 (0.67, 1.11) | 0.25 |
| Lysine | 0.85 (0.8, 0.91) | < **0.001** |
| Shikimic acid | 0.85 (0.74, 0.99) | **0.032** |
| Histidine | 0.84 (0.79, 0.88) | < **0.001** |
| Adipic acid | 0.84 (0.72, 0.96) | **0.014** |
| TMAO | 0.84 (0.6, 1.16) | 0.29 |
| Glycine | 0.83 (0.77, 0.9) | < **0.001** |
| Pentothenate | 0.83 (0.73, 0.94) | **0.0036** |
| Oxaloacetate | 0.81 (0.74, 0.89) | < **0.001** |
| GDP | 0.8 (0.74, 0.88) | < **0.001** |
| Tryptophan | 0.78 (0.73, 0.84) | < **0.001** |
| Aconitate | 0.78 (0.72, 0.84) | < **0.001** |
| Glyceraldehyde | 0.78 (0.69, 0.9) | < **0.001** |
| Oxypurinol | 0.78 (0.67, 0.9) | < **0.001** |

| | | |
|---|---|---|
| Proline | 0.77 (0.72, 0.83) | < **0.001** |
| Asparagine | 0.75 (0.71, 0.79) | < **0.001** |
| Arginine | 0.75 (0.66, 0.85) | < **0.001** |
| Valine | 0.74 (0.69, 0.8) | < **0.001** |
| Ornithine | 0.74 (0.68, 0.82) | < **0.001** |
| Threonine | 0.74 (0.68, 0.8) | < **0.001** |
| Phenylalanine | 0.73 (0.69, 0.77) | < **0.001** |
| Guanidinoacetate | 0.73 (0.66, 0.8) | < **0.001** |
| N-AcetylGlycine | 0.73 (0.64, 0.83) | < **0.001** |
| Hydroxyproline | 0.73 (0.61, 0.88) | < **0.001** |
| MethylSuccinate | 0.72 (0.67, 0.78) | < **0.001** |
| Serine | 0.71 (0.66, 0.76) | < **0.001** |
| IMP | 0.71 (0.47, 1.07) | 0.098 |
| 2-Hydroxyglutarate | 0.7 (0.62, 0.78) | < **0.001** |
| Glutamic acid | 0.7 (0.6, 0.82) | < **0.001** |
| Cystathionine | 0.7 (0.59, 0.84) | < **0.001** |
| Methionine | 0.67 (0.62, 0.71) | < **0.001** |
| Sorbitol | 0.66 (0.6, 0.72) | < **0.001** |
| Tyrosine | 0.65 (0.61, 0.69) | < **0.001** |
| Succinate | 0.61 (0.55, 0.68) | < **0.001** |
| PGE | 0.6 (0.53, 0.69) | < **0.001** |
| Citrulline | 0.58 (0.53, 0.62) | < **0.001** |
| Leucine | 0.56 (0.51, 0.61) | < **0.001** |
| iso-Leucine | 0.55 (0.5, 0.6) | < **0.001** |
| Adenylosuccinate | 0.53 (0.44, 0.65) | < **0.001** |
| Aspartic acid | 0.51 (0.43, 0.6) | < **0.001** |
| Taurine | 0.49 (0.41, 0.58) | < **0.001** |
| Malonic acid | 0.38 (0.28, 0.51) | < **0.001** |
| Niacinamide | 0.37 (0.29, 0.48) | < **0.001** |
| ADP | 0.21 (0.14, 0.32) | < **0.001** |
| Linolenic acid | 0.18 (0.15, 0.21) | < **0.001** |

**Supplemental table 2.** Differences in plasma metabolic response post glucose challenge by CKD status. Result from regression analysis using SERFF normalized on sample type (fasting vs OGTT) by CKD status adjusted for age, sex, race, weight, and batch. Fold changes represent changes in metabolite levels with fasting levels after glucose load. (e.g. a fold change of 0.92 indicates a reduction of %8 in metabolite level. P-value for interaction represents the heterogeneity in fold change by disease status.

| Metabolite name | Fold change between OGTT/fasting for Non-CKD (%95 Cl) | Fold change between OGTT/fasting for CKD (%95 Cl) | p-value for interaction |
|---|---|---|---|
| Succinate | 0.48 (0.37, 0.62) | 0.78 (0.71, 0.85) | < 0.001 |
| Taurine | 0.41 (0.28, 0.6) | 0.75 (0.66, 0.85) | 0.0032 |
| Adenylosuccinate | 0.41 (0.26, 0.65) | 0.84 (0.72, 0.98) | 0.0037 |
| Hippuric acid | 1.56 (0.67, 3.59) | 5.68 (4.29, 7.54) | 0.004 |
| ADP | 0.17 (0.07, 0.42) | 0.63 (0.47, 0.86) | 0.0063 |
| Biotin | 0.74 (0.59, 0.93) | 1.02 (0.94, 1.1) | 0.011 |
| Niacinamide | 0.35 (0.2, 0.6) | 0.7 (0.58, 0.84) | 0.019 |
| Glycochenodeoxycholate | 0.47 (0.16, 1.35) | 1.69 (1.18, 2.42) | 0.025 |
| Inositol | 0.92 (0.85, 0.99) | 1 (0.98, 1.03) | 0.026 |
| Kynurenate | 1.07 (0.82, 1.4) | 0.78 (0.71, 0.85) | 0.026 |
| IMP | 0.18 (0.07, 0.48) | 0.55 (0.39, 0.76) | 0.037 |
| Uridine | 0.69 (0.49, 0.96) | 0.99 (0.89, 1.11) | 0.043 |
| Glucose | 1.07 (0.88, 1.31) | 1.34 (1.25, 1.43) | 0.043 |
| GDP | 0.73 (0.59, 0.89) | 0.9 (0.84, 0.97) | 0.05 |
| Melatonin | 0.48 (0.28, 0.84) | 0.83 (0.69, 1) | 0.067 |
| Glycocholate | 0.7 (0.29, 1.66) | 1.61 (1.2, 2.15) | 0.073 |
| 2-Hydroxyglutarate | 0.57 (0.43, 0.76) | 0.73 (0.67, 0.8) | 0.1 |
| iso-Leucine | 0.49 (0.4, 0.59) | 0.58 (0.54, 0.61) | 0.12 |
| Urate | 1.06 (1.02, 1.1) | 1.03 (1.01, 1.04) | 0.12 |
| D-Leucic.Acid | 0.68 (0.44, 1.04) | 0.96 (0.83, 1.11) | 0.13 |
| Creatinine | 0.87 (0.78, 0.97) | 0.95 (0.91, 0.98) | 0.15 |
| Aspartic.Acid | 0.63 (0.47, 0.83) | 0.78 (0.71, 0.86) | 0.15 |
| 1-Methyladenosine | 0.89 (0.81, 0.99) | 0.97 (0.94, 1.01) | 0.15 |
| Guanidinoacetate | 0.67 (0.54, 0.82) | 0.78 (0.73, 0.83) | 0.15 |
| Pentothenate | 0.72 (0.54, 0.96) | 0.9 (0.81, 0.99) | 0.17 |
| Glycerate | 0.75 (0.5, 1.11) | 0.99 (0.87, 1.14) | 0.18 |

| | | | |
|---|---|---|---|
| Aconitate | 0.83 (0.69, 1) | 0.95 (0.9, 1.02) | 0.18 |
| Pyridoxal-5-P | 0.82 (0.56, 1.18) | 1.07 (0.94, 1.21) | 0.18 |
| Trimethylamine-N-oxide.(TMAO) | 0.41 (0.19, 0.87) | 0.69 (0.54, 0.9) | 0.2 |
| F16BP/F26BP/G16BP | 0.92 (0.84, 1.02) | 0.99 (0.96, 1.02) | 0.2 |
| Citraconic.Acid | 1.29 (0.78, 2.13) | 0.92 (0.78, 1.09) | 0.21 |
| Erythrose | 1.45 (0.89, 2.37) | 1.04 (0.89, 1.23) | 0.21 |
| Glutamic.acid | 0.59 (0.44, 0.79) | 0.72 (0.65, 0.79) | 0.22 |
| Leucine | 0.51 (0.42, 0.62) | 0.57 (0.54, 0.61) | 0.28 |
| Glutaric.Acid/Oxaloacetate | 0.87 (0.72, 1.05) | 0.97 (0.91, 1.03) | 0.28 |
| Aminoisobutyrate | 1.04 (0.83, 1.29) | 0.92 (0.86, 0.99) | 0.32 |
| Proline | 0.77 (0.66, 0.9) | 0.84 (0.79, 0.88) | 0.32 |
| Hydroxyproline/Aminolevulinate | 0.64 (0.42, 0.98) | 0.79 (0.69, 0.92) | 0.34 |
| Pyruvate | 1.08 (0.71, 1.64) | 1.34 (1.16, 1.54) | 0.34 |
| Serine | 0.75 (0.64, 0.89) | 0.82 (0.77, 0.87) | 0.37 |
| Cystamine | 1.09 (0.74, 1.6) | 0.9 (0.79, 1.03) | 0.38 |
| L-Kynurenine | 1.09 (0.9, 1.32) | 0.99 (0.93, 1.06) | 0.38 |
| Arginine | 0.72 (0.53, 0.99) | 0.84 (0.75, 0.93) | 0.39 |
| Adipic.Acid | 1.04 (0.75, 1.43) | 0.89 (0.8, 0.99) | 0.39 |
| Valine | 0.7 (0.61, 0.82) | 0.75 (0.72, 0.79) | 0.4 |
| Carnitine | 0.97 (0.84, 1.11) | 1.02 (0.98, 1.07) | 0.42 |
| Phenylalanine | 0.74 (0.66, 0.84) | 0.78 (0.75, 0.81) | 0.42 |
| Asparagine | 0.75 (0.67, 0.84) | 0.79 (0.76, 0.82) | 0.44 |
| Glyceraldehyde | 0.8 (0.59, 1.07) | 0.9 (0.81, 1) | 0.44 |
| Chenodeoxycholate | 0.94 (0.79, 1.12) | 0.88 (0.82, 0.93) | 0.45 |
| Citrulline | 0.63 (0.53, 0.74) | 0.67 (0.63, 0.71) | 0.48 |
| Shikimic.Acid | 1.17 (0.89, 1.55) | 1.06 (0.96, 1.16) | 0.49 |
| Hypoxanthine | 0.92 (0.7, 1.21) | 1.01 (0.92, 1.11) | 0.52 |
| Linolenic.Acid | 0.23 (0.17, 0.32) | 0.26 (0.23, 0.29) | 0.52 |
| 5-Hydroxytryptophan | 1.06 (0.97, 1.15) | 1.03 (1, 1.06) | 0.54 |
| GTP | 0.95 (0.86, 1.05) | 0.98 (0.95, 1.02) | 0.54 |
| Glycine | 0.8 (0.67, 0.94) | 0.84 (0.79, 0.89) | 0.56 |
| Oxypurinol | 0.63 (0.44, 0.9) | 0.7 (0.62, 0.79) | 0.6 |
| Xanthine | 0.92 (0.71, 1.19) | 0.99 (0.9, 1.08) | 0.61 |
| Cystathionine | 0.9 (0.66, 1.23) | 0.82 (0.74, 0.92) | 0.61 |

| | | | |
|---|---|---|---|
| Dimethylglycine | 0.79 (0.58, 1.06) | 0.85 (0.77, 0.95) | 0.62 |
| Glutamine | 0.92 (0.77, 1.09) | 0.88 (0.83, 0.93) | 0.62 |
| N-AcetylGlycine | 0.71 (0.52, 0.96) | 0.77 (0.69, 0.85) | 0.62 |
| Betaine | 1.01 (0.84, 1.2) | 0.96 (0.9, 1.02) | 0.63 |
| Tyrosine | 0.68 (0.59, 0.78) | 0.7 (0.67, 0.74) | 0.63 |
| Lysine | 0.83 (0.72, 0.96) | 0.86 (0.82, 0.9) | 0.64 |
| Oxalic.Acid | 1.26 (0.91, 1.74) | 1.16 (1.04, 1.29) | 0.64 |
| Methionine | 0.67 (0.58, 0.78) | 0.7 (0.66, 0.73) | 0.65 |
| Alanine | 0.89 (0.72, 1.11) | 0.94 (0.87, 1.01) | 0.67 |
| Threonine | 0.78 (0.67, 0.91) | 0.75 (0.72, 0.79) | 0.69 |
| Malonic Acid | 0.95 (0.63, 1.43) | 1.04 (0.9, 1.19) | 0.69 |
| Histidine | 0.86 (0.76, 0.96) | 0.84 (0.8, 0.87) | 0.7 |
| MethylSuccinate | 0.76 (0.64, 0.89) | 0.78 (0.74, 0.82) | 0.73 |
| Homovanilate | 0.98 (0.74, 1.29) | 0.93 (0.85, 1.02) | 0.73 |
| Kuraridinol | 1.07 (0.84, 1.37) | 1.02 (0.95, 1.1) | 0.74 |
| Creatine | 1.14 (0.85, 1.53) | 1.08 (0.98, 1.2) | 0.75 |
| Trimethylamine (TMA) | 1.04 (0.9, 1.21) | 1.06 (1.01, 1.12) | 0.78 |
| Propionate | 0.83 (0.58, 1.19) | 0.79 (0.7, 0.89) | 0.78 |
| Sorbitol | 0.67 (0.55, 0.8) | 0.68 (0.64, 0.73) | 0.8 |
| lactate | 1.15 (0.82, 1.61) | 1.2 (1.08, 1.35) | 0.81 |
| Allantoin | 0.93 (0.76, 1.14) | 0.95 (0.89, 1.02) | 0.81 |
| Choline | 0.98 (0.78, 1.22) | 0.95 (0.88, 1.03) | 0.83 |
| Fumaric.Acid/Maleic.Acid | 0.95 (0.77, 1.19) | 0.94 (0.87, 1.01) | 0.89 |
| PGE | 0.75 (0.59, 0.96) | 0.77 (0.71, 0.83) | 0.89 |
| Glycochenodeoxycholic.acid | 0.83 (0.07, 9.83) | 0.7 (0.34, 1.44) | 0.9 |
| Ornithine | 0.77 (0.62, 0.97) | 0.76 (0.71, 0.82) | 0.92 |
| G1P/G6P/F6P/F1P | 0.95 (0.85, 1.06) | 0.95 (0.91, 0.98) | 0.94 |
| Glucoronate | 0.92 (0.7, 1.22) | 0.92 (0.84, 1.01) | 0.98 |
| Cystine | 0.98 (0.83, 1.14) | 0.98 (0.93, 1.03) | 0.99 |
| Tryptophan | 0.86 (0.73, 1.01) | 0.86 (0.81, 0.91) | 1 |

**Supplemental table 3.** The list of metabolites in each module from the WGCNA analysis.

| Module color | Metabolite name |
|---|---|
| black | Alanine |
| black | Pyruvate |
| black | lactate |
| black | Oxalic.Acid |
| blue | Glycine |
| blue | Aminoisobutyrate |
| blue | Choline |
| blue | Serine |
| blue | Asparagine |
| blue | Phenylalanine |
| blue | Arginine |
| blue | MethylSuccinate |
| blue | Citrulline |
| blue | G1P.G6P.F6P.F1P |
| blue | Prostaglandin E |
| brown | TMAO |
| brown | Dimethylglycine |
| brown | Proline |
| brown | Threonine |
| brown | Hydroxyproline |
| brown | Glutamine |
| brown | Propionate |
| brown | Hyppuric acid |
| brown | Linolenic acid |
| green | Malonic acid |
| green | D-Leucic acid |
| green | Urate |
| green | Inositol |
| green | Kynurenate |
| green | IMP |
| grey | Trimethylamine |

| | |
|---|---|
| grey | Creatinine |
| grey | Betaine |
| grey | Creatine |
| grey | Glutamic acid |
| grey | Carnitine |
| grey | 13C-Arginine |
| grey | 13C-Tyrosine |
| grey | L-Kynurenine |
| grey | 5-Hydroxytryptophan |
| grey | Cystine |
| grey | Uridine |
| grey | 1-Methyladenosine |
| grey | Glyceraldehyde |
| grey | C13-Lactate |
| grey | Glycerate |
| grey | N-AcetylGlycine |
| grey | Citraconic acid |
| grey | Hypoxanthine |
| grey | Adipic acid |
| grey | 2-Hydroxyglutarate |
| grey | Oxypurinol |
| grey | Xanthine |
| grey | Allantoin |
| grey | Shikimic acid |
| grey | Aconitate |
| grey | Glucose |
| grey | 13C-Glucose |
| grey | Glucoronate |
| grey | Pentothenate |
| grey | Cystathionine |
| grey | Melatonin |
| grey | Biotin |
| grey | Pyridoxal-5-P |
| grey | F16BP.F26BP.G16BP |

| | |
|---|---|
| grey | Chenodeoxycholate |
| grey | Glycochenodeoxycholate |
| grey | Glycocholate |
| grey | GTP |
| red | Fumaric acid |
| red | Glutaric acid |
| red | Homovanilate |
| red | Erythrose |
| turquoise | Valine |
| turquoise | Leucine |
| turquoise | Iso-Leucine |
| turquoise | Ornithine |
| turquoise | Lysine |
| turquoise | Methionine |
| turquoise | Cystamine |
| turquoise | Histidine |
| turquoise | Tyrosine |
| turquoise | Sorbitol |
| turquoise | Tryptophan |
| turquoise | Guanidinoacetate |
| yellow | Niacinamide |
| yellow | Taurine |
| yellow | Aspartic acid |
| yellow | Succinate |
| yellow | ADP |
| yellow | GDP |
| yellow | Adenylosuccinate |
